# Characterizing resting-state EEG oscillatory and aperiodic activity in neurodegenerative diseases: A multicentric study

**DOI:** 10.1101/2025.02.14.25322283

**Authors:** Alberto Jaramillo-Jimenez, Yorguin-Jose Mantilla-Ramos, Diego A Tovar-Rios, Francisco Lopera, David Aguillón, John Fredy Ochoa-Gomez, Claire Paquet, Sinead Gaubert, Matteo Pardini, Dario Arnaldi, John-Paul Taylor, Tormod Fladby, Kolbjørn Brønnick, Dag Aarsland, Laura Bonanni, the E-DLB Consortium

## Abstract

**Background:** Abnormalities in resting-state electroencephalogram (rsEEG) posterior alpha rhythm are promising biomarkers of neurodegenerative diseases (NDDs), often assessed via spectral analysis, ignoring the signal’s non-rhythmic (aperiodic) component. Evidence assessing aperiodic and oscillatory rsEEG abnormalities across NDDs is scarce and often underpowered. Multicenter studies could tackle these limitations, but data pooling might introduce site-related rsEEG differences (batch effects). This study aims to characterize rsEEG oscillatory and aperiodic patterns across NDDs, minimizing potential batch effects.

**Methods:** RsEEGs (n = 639; 11 sites) were automatically preprocessed. Signals comprised healthy controls (HC = 153), Lewy Body Dementias (LBD = 95), Parkinson’s Disease (PD = 71), Alzheimer’s Disease (AD = 186), Frontotemporal Dementia (FTD = 23), Mild Cognitive Impairment (MCI) in positive Lewy Body pathology or PD (MCI-LBD = 34), and MCI in positive AD pathology (MCI-AD = 77). Power spectrum batch effects were harmonized using reComBat (age, sex, and diagnosis-adjusted). Harmonization was evaluated with functional and mass-univariate ANOVAs. Oscillatory and aperiodic parameters were extracted from the batch-harmonized power spectrum. NDDs-related differences were estimated with functional and mass-univariate tests, bootstrapped pairwise comparisons, and logistic regressions.

**Results:** Qualitative visualizations and statistical testing showed reduced batch effects after harmonization. Statistically significant findings included steeper aperiodic parameters and lower oscillatory center frequency in LBD compared to other NDDs. Additionally, oscillatory extended alpha power was lower in AD vs. LBD.

**Conclusions:** Batch effects in the rsEEG power spectrum can be mitigated with harmonization. Oscillatory alpha power reduction may better reflect AD abnormalities, whereas pronounced oscillatory frequency slowing and greater aperiodic activity characterize LBD.

## Introduction

Since Hans Berger’s pioneering reports on the human electroencephalogram, The reduction in posterior dominant alpha rhythm has been observed in both physiological and pathological aging, being particularly evident in dementia patients (Berger, 1933). Beyond qualitative descriptors, quantitative analysis of the resting-state electroencephalogram (rsEEG) paved the way for its broader application in dementia research (Al-Qazzaz et al., 2014). The “slowing” of posterior dominant rsEEG rhythms from alpha (8 – 13 Hz) to theta frequencies (4 – 8 Hz), has been consistently reported in multiple neurodegenerative diseases (NDDs) that lead to dementia syndrome, including Alzheimer’s Disease (AD), Parkinson’s Disease (PD), Dementia with Lewy Bodies (DLB), and Frontotemporal Dementia (FTD) (Babiloni et al., 2020b; Dauwels et al., 2011; Dringenberg, 2000; Eichelberger et al., 2017; Franciotti et al., 2020; Zimmermann et al., 2015). As a case in point, posterior alpha peak frequency slowing, with or without increased variability, is included as a supportive biomarker in the current DLB diagnostic criteria (McKeith et al., 2017), whereas reductions in alpha power and peak frequency have been recommended by expert panels as candidate features for diagnosis, treatment monitoring, and prognosis in the AD continuum (Babiloni et al., 2021).

Despite this promising background, the methodological nuances in the analysis of rsEEG studies often limit the generalization of their results. Differences in analysis pipelines across research laboratories preclude direct comparability of their statistical estimations via secondary studies (e.g., meta-analysis) (Bigdely-Shamlo et al., 2020). Furthermore, the small sample sizes typical of most single-site rsEEG studies undermine both statistical power and external validity (Button et al., 2013; Larson and Carbine, 2017; Newson and Thiagarajan, 2019). Multicentric collaborations aim to address these limitations by aggregating data from various research sites, resulting in larger and more heterogeneous datasets (Bonanni et al., 2016; Li et al., 2022; Prado et al., 2023; Thompson et al., 2014). Although simple pooling of single-site data increases the sample size, insights from recent multicenter initiatives highlight the importance of assessing and mitigating batch effects (i.e., non-biological, systematic cross-site differences in rsEEG data distributions that can confound the results) (Bayer et al., 2022; Hu et al., 2023; Li et al., 2022; Marzi et al., 2024).

Batch effects stem from technical factors, including distinct scanners (headsets/amplifiers) or acquisition parameters (Bigdely-Shamlo et al., 2020; Li et al., 2022). Besides, differences in the rsEEG data distributions could arise from biological characteristics of the pooled sample (such as site-specific age distributions) (Bayer et al., 2022; Hu et al., 2023; Li et al., 2022). Among the statistical strategies to mitigate batch effects while preserving the effects of biological covariates of interest (Bayer et al., 2022; Fortin et al., 2018; Hu et al., 2023), the Combining Batches (ComBat) method has gained popularity in genetics (Adamer et al., 2022), proteomics (Voß et al., 2022), and neuroimaging (Bell et al., 2022; Fortin et al., 2018, 2017; Horng et al., 2022a, 2022b; Pomponio et al., 2020), with numerous validations and adaptations improving the original ComBat algorithm (initially formulated for microarray expression data) (Johnson et al., 2007). Nonetheless, evidence evaluating ComBat-derived methods to harmonize batch effects on rsEEG-derivative metrics is scarce (Jaramillo-Jimenez et al., 2024; Li et al., 2022).

In earlier publications (Bonanni et al., 2016; Franciotti et al., 2020), our group pooled datasets from individual research centers to assess rsEEG spectral patterns in DLB (i.e., remarkable posterior peak frequency shifting and variability) exhibiting consistent results with single-site observations (Law et al., 2020; Massa et al., 2020; Schumacher et al., 2020; van der Zande et al., 2018). Nevertheless, these prior works did not account for batch effects assessment and correction.

On the other hand, most of our preliminary analyses in DLB used the rsEEG power spectrum to represent brain rhythms without modeling the contribution of non-rhythmic activity (so-called 1/frequency, scale-free, fractal, aperiodic, or non-oscillatory). Recent models propose that the neuronal power spectrum displays a mixture of aperiodic activity (observed as a power slope that decreases as frequency increases) with superimposed oscillatory peaks (Brake et al., 2024). Moreover, results from simulations and real rsEEG data suggest that descriptors of the power spectrum (such as absolute and relative power or peak frequency) could be conflated due to the effect of the aperiodic activity obscuring real oscillatory differences (Donoghue et al., 2020; Gerster et al., 2022). Parameterizing aperiodic activity is also supported by its neurophysiological relevance (Brake et al., 2024), with potential applications in neurodevelopment and age-related medicine (Donoghue et al., 2020; McSweeney et al., 2023; Merkin et al., 2023; Voytek et al., 2015), anesthesia (Colombo et al., 2019), and sleep (Bódizs et al., 2024). Compared to rsEEG oscillatory descriptors, the characterization of aperiodic activity has not been extensively assessed across the different clinical phenotypes of NDDs, except by recent reports in small samples suggesting the hypotheses of predominantly oscillatory abnormalities in AD (Kopčanová et al., 2024) and aperiodic changes in PD and DLB (McKeown et al., 2023; Rosenblum et al., 2023; Wang et al., 2024).

In light of the above, this study aims to: A) characterize rsEEG oscillatory and aperiodic patterns across multiple clinical phenotypes of NDDs, and B) evaluate and mitigate potential batch effects in multicentric rsEEG data. Through a standardized preprocessing and analysis workflow, we provide an open pipeline that can be extended for group-level analysis of other multicentric rsEEG datasets.

## Materials and methods

### Study Design and Settings

This secondary analysis capitalized on data from cross-sectional individual studies (n = 11) conducted in eight countries (Colombia, Finland, France, Greece, Italy, Norway, the United Kingdom, and the United States) to assess rsEEG biomarkers in NDDs and healthy aging.

Beyond openly available (open) datasets (Anjum et al., 2020; Hatlestad-Hall, 2022; Miltiadous et al., 2023; Railo, 2021; Rockhill et al., 2021), we used in-house clinical research data collected cross-sectionally by the European Dementia with Lewy Bodies Consortium (EDLB) (Oppedal et al., 2019), the Dementia Disease Initiation Study (DDI) (Fladby et al., 2017), and Grupo de Neurociencias de Antioquia (GNA) (Carmona Arroyave et al., 2019; Jaramillo-Jimenez et al., 2021).

Briefly, datasets from the following locations were included: California, United States (open); Chieti, Italy (EDLB); Genoa, Italy (EDLB); Iowa, United States (open); Medellin, Colombia (GNA); Newcastle, United Kingdom (EDLB); Oslo, Norway (open); Paris, France (EDLB); Stavanger, Norway (EDLB); Stavanger, Norway (DDI); Thessaloniki, Greece (open) and Turku, Finland (open). Supplementary Table 1 provides an extended description of locations and sources of data.

### Participants

The pooled sample (n = 639) comprised rsEEG signals from 153 healthy controls (HC) and 486 individuals with a clinical diagnosis of NDD. Phenotypes of early and late-onset neurodegenerative causes of dementia were included, such as probable AD, FTD, DLB, and dementia in PD (PDD). Additionally, subgroups comprised mild cognitive impairment (MCI) in the context of different NDDs: positive Lewy Body pathology (MCI-LB), Parkinson’s disease (MCI-PD), and positive Alzheimer’s disease-related pathology (MCI-AD). These, in conjunction with an additional cognitively normal Parkinson’s disease group (PD), served to appraise the continuum of predementia stages. Detailed information on clinical criteria used for diagnosis is presented in Supplementary Table 2. In all the primary data sources, neuropsychiatric diseases other than the abovementioned NDDs were excluded. All individuals provided their informed consent prior to recruitment in the primary studies. All primary studies were approved by their local, institutional, or regional ethics committees preserving the World Medical Association Declaration of Helsinki.

Given preliminary evidence indicating shared pathological substrates in PDD and DLB (Jellinger and Korczyn, 2018), we merged these subgroups into a Lewy Body Dementia (LBD) group. In line with this, we combined the MCI-LB and MCI-PD subgroups, henceforth referring to them collectively as MCI-LBD.

In the Oslo, Norway dataset (with young and old healthy individuals), a subsample of comparable HC subjects was retained for subsequent analyses based on the minimum age value across subjects with NDDs. Thus, only healthy participants aged 44 or older were included, excluding younger individuals.

### RsEEG acquisition and signal preprocessing

The raw rsEEG data from open datasets is publicly available in online repositories (Anjum et al., 2020; Hatlestad-Hall, 2022; Miltiadous et al., 2023; Railo, 2021; Rockhill et al., 2021).

Acquisition parameters varied broadly across primary studies. RsEEG signals were recorded during the eyes closed condition in all datasets except data from Iowa, USA, which was recorded under the eyes open condition. The cross-study differences in the number of channels, sampling frequency, amplifier/headset, and recording length are presented in Supplementary Table 3.

To achieve comparability, signals were down-sampled to a common sample frequency of 128 Hz (ranging from 128 – 1024 Hz). Electrodes were placed and named following the international 10-20, 10-10, and 10-05 system distributions (Seeck et al., 2017), and the number of electrodes ranged from 19 to 128 leads across sites. Therefore, 18 common rsEEG channels placed accordingly to the international 10-05 system were preserved for further analysis [‘Fp1’, ‘Fp2’, ‘F3’, ‘F4’, ‘F7’, ‘F8’, ‘Fz’, ‘C3’, ‘C4’, ‘Cz’, ‘T7’, ‘T8’, ‘P3’, ‘P4’, ‘P7’, ‘P8’, ‘O1’, ‘O2’].

All primary datasets were standardized following the Brain Image Data Structure (BIDS) specification (Pernet et al., 2019) and automatically preprocessed using sovaharmony (available at https://github.com/GRUNECO/eeg_harmonization), validated elsewhere by our group (Isaza et al., 2023; Jaramillo-Jimenez et al., 2023, 2021; Suarez-Revelo et al., 2016, 2018). Briefly, our preprocessing workflow is a wrapper of multiple utilities. First, to obtain a comparable reference scheme across studies without the influence of noisy channels, robust average re-referencing, adaptative line-noise correction, and bad channel interpolation were conducted using the PyPREP library (Appelhoff et al., 2022; Bigdely-Shamlo et al., 2015). Subsequently, a 1 Hz high-pass Finite Impulse Response filter was used to remove low-frequency drifts before applying wavelet-enhanced independent component analysis (ICA) artifact smoothing (Castellanos and Makarov, 2006). For this purpose, the FastICA algorithm implemented in the MNE library (Gramfort et al., 2013) was applied to obtain both artifactual and brain components; then wavelet thresholding was used to subtract strong muscular and eye-blink components. Next, signals were low-pass filtered at 45 Hz and segmented into five-second-length epochs (5 s). Finally, artifactual epoch rejection was conducted based on signal parameters (e.g. extreme amplitude and spectral power values) and statistical properties (e.g. linear trends, joint probability, and kurtosis).

The number of non-artifactual epochs varied across studies depending on each specific protocol. For this study, only signals with a length greater or equal to 100 seconds (20 epochs) after preprocessing were included, following preliminary evidence supporting good test-retest reliability of spectral rsEEG features in healthy (Gudmundsson et al., 2007) and DLB subjects (Jin et al., 2023) with more than 60 – 90 seconds length. Supplementary Figure 1 presents the positions of included electrodes and available non-artifactual epochs across sites.

### Feature Extraction and Batch Harmonization

Before feature extraction, we equalized the number of epochs across all subjects selecting the 20 epochs with the highest quality based on the scorEpochs algorithm (Fraschini et al., 2022), available at https://github.com/Scorepochs-tools/scorepochs_py. ScorEpochs computes a similarity score (Spearman correlation coefficient) from the power spectrum (computed with the Welch method) at the channel and epochs level, assessing the quality of each epoch. Further, power spectrum vectors were obtained at the sensor level using the psd_array_multitaper function implemented in MNE Python with default parameters and a frequency range from 1 – 30 Hz. This frequency range was chosen based on a systematic review showing that 50% of the clinical studies estimating aperiodic activity used a range of 1-43 Hz and 85% have adopted the 3–30 Hz range (Donoghue, 2024). Then, the median power spectrum across the 20 epochs was computed, returning a single power spectrum vector per channel. As this study targeted the dominant alpha rhythm, a median power spectrum vector across channels in the posterior region of interest [’P3’, ‘P4’, ‘P7’, ‘P8’, ‘O1’, ‘O2’] was calculated per subject.

Batch harmonization of the median posterior power spectrum vectors was performed using the reComBat algorithm version 0.1.4 (Adamer et al., 2022), available at https://github.com/BorgwardtLab/reComBat. ReComBat assumes that the harmonized features can be modeled as a linear combination of the site-related batch effects, biological covariates, and a site-specific error term. Extending other versions of ComBat (Fortin et al., 2017; Horng et al., 2022a; Johnson et al., 2007; Pomponio et al., 2020; Voß et al., 2022), reComBat mitigates batch effects from the extracted features by modeling site-specific scaling factors while preserving biological covariates even in cases of singular design matrixes (e.g., subjects from the same site with a single unique diagnosis). To solve the singular design covariates matrix, reComBat implements Lasso, Ridge, or Elastic-net regularization. A graphical representation of the reComBat model is presented in Supplementary Figure 2. In this study, we fitted a parametric reComBat model with Elastic-net regularization (alpha=1e-6; max_iter=50000) to harmonize the posterior power spectrum across sites adjusting for sex, age, and diagnosis effects.

### Spectral Parameterization

Given the potential confounder effects of aperiodic activity in the median posterior power spectrum vectors, spectral parameterization was conducted using the Fitting Oscillations & One Over Frequency algorithm (FOOOF) version 1.1 (Donoghue et al., 2020), available at https://fooof-tools.github.io/fooof/. The FOOOF method has been successfully applied to study age-related rsEEG aperiodic changes in healthy and diseased populations (Kopčanová et al., 2024; McKeown et al., 2023; Merkin et al., 2023; Rosenblum et al., 2023; Wang et al., 2024) with good test-retest reliability (McKeown et al., 2024; Popov et al., 2023).

The power spectrum is an admixture of neuronal oscillations (peaks) superimposed on non-oscillatory (aperiodic) activity which follows a 1/frequency distribution (i.e., higher power values at slow frequencies that decrease as frequency increases). Thus, spectral parameterization decomposes the oscillatory and non-oscillatory activity from the power spectrum. First, the FOOOF algorithm takes an input power spectrum in the linear scale and runs a linear fit to the aperiodic trend in the log-log scale with a Lorentzian function. This aperiodic fit is then subtracted from the input power spectrum, “exposing” the oscillatory peaks. Oscillatory peaks are subsequently modeled through iterative Gaussian fits. Finally, FOOOF returns the fitted aperiodic and oscillatory activity vectors. An extensive review is available elsewhere (Donoghue et al., 2020; Gerster et al., 2022). In this study, the FOOOF model was fitted between 1 – 30 Hz using the following parameters (peak_width_limits=[1, 8], min_peak_height=0.05, max_n_peaks=6).

From the derivative vectors (i.e., isolated oscillatory and aperiodic activity), FOOOF computes the following descriptors: Oscillatory parameters (Power – PW, Bandwidth – BW, Center Frequency – CF), Aperiodic parameters (Exponent, Offset), Fitting parameters (error and R-squared). For the present study, we estimated oscillatory parameters in the extended alpha band (5 – 14 Hz) (Moretti et al., 2013). Similarly, the aperiodic exponent (representing the slope estimated from the fitted 1/frequency activity) and the aperiodic offset (representing the y-axis intercept of the 1/frequency activity) were also estimated. Only subjects with good quality of FOOOF fitting were included (i.e., Model’s R-squared greater or equal to 0.8). Supplementary Figure 3 illustrates the spectral parameterization fitting with FOOOF, and the derivative vectors and parameters.

### Statistical Analysis

All statistical analyses were performed in Python (v. 3.10.15), using the scikit-fda (v. 0.9.1), MNE Python (v. 1.5.1), and pingouin (v. 0.5.5) libraries (Cuevas et al., 2004; Gramfort et al., 2013; Vallat, 2018). The alpha significance level was set below 0.05 (multiple-testing corrected).

Demographic characteristics of the samples were presented with descriptive statistics. Welch ANOVA and Chi-squared tests were performed to explore potential age, sex, and diagnosis differences across sites.

Following previously published recommendations on batch effects harmonization (Hu et al., 2023), we evaluated potential batch and harmonization effects using qualitative visualizations and statistical testing across batches. Specifically, bivariate plots (power vs. frequency) and Principal Component Analysis (PCA) plots were generated from the unharmonized and batch-harmonized power spectrum. PCA reduces the dimensionality of the power spectrum (146 values across all frequency bins for each subject) into a lower space, maximizing the variance across subjects. The Euclidian distances between PCA site centroids indicated batch effects (pairwise and cross-sites). Further, functional and mass-univariate ANOVAs were used to quantify batch effects before and after harmonization. Functional ANOVA (Cuevas et al., 2004) applies principles from Functional Data Analysis (Jaramillo-Jimenez et al., 2023; Ramsay, 2012; Tian, 2010; Ullah and Finch, 2013) to model the entire power spectrum of each subject as a single functional observation (dependent variable) rather than discrete points. This approach estimates the differences across sites (independent variable) based on the ANOVA design, quantifying batch and harmonization effects in the power spectrum as continuous functions. Functional ANOVA was performed using the function oneway_anova (unequal variance, with b-spline basis order = 4, number of bases = 14) from the scikit-fda utility available at https://github.com/GAA-UAM/scikit-fda. Complementarily, site-related differences were estimated across each frequency bin of the power spectrum using mass-univariate ANOVAs. Multiple testing was addressed with permutation tests clustered on frequencies as implemented in MNE Python’s function permutation_cluster_test (n_permutations = 2000, two tails).

NDDs-related differences were estimated in the batch-harmonized power spectrum as well as the derivative oscillatory and activity. Functional and mass-univariate ANOVAs explored differences in the power spectrum across all NDDs (independent variable). To determine specific frequencies with distinctive rsEEG activity, pairwise differences across diagnosis subgroups were estimated on the batch-harmonized power spectrum and on the derivative oscillatory and aperiodic-fitted vectors using mass-univariate permutation cluster tests (n_permutations = 1000, two tails). Complementarily, differences in the extended alpha oscillatory parameters (CF, PW, BW), and the broadband aperiodic parameters (exponent, offset) were assessed through pairwise comparisons. Thus, t-tests with bootstrapped confidence intervals (1000 iterations) were performed across diagnosis subgroups utilizing the pairwise_tests and compute_bootci implemented in Python’s statistical library: pingouin. Multiple testing was addressed with the Benjamini-Yekutieli False Discovery Ratio correction method (Benjamini and Yekutieli, 2001). Finally, separate multinomial logistic regression models were fitted for each oscillatory and aperiodic parameter (independent variable) to estimate age– and sex-adjusted associations with diagnosis subgroups (dependent variable). A graphical abstract summarizing the study methods is presented in Figure 1.

**Figure 1.**
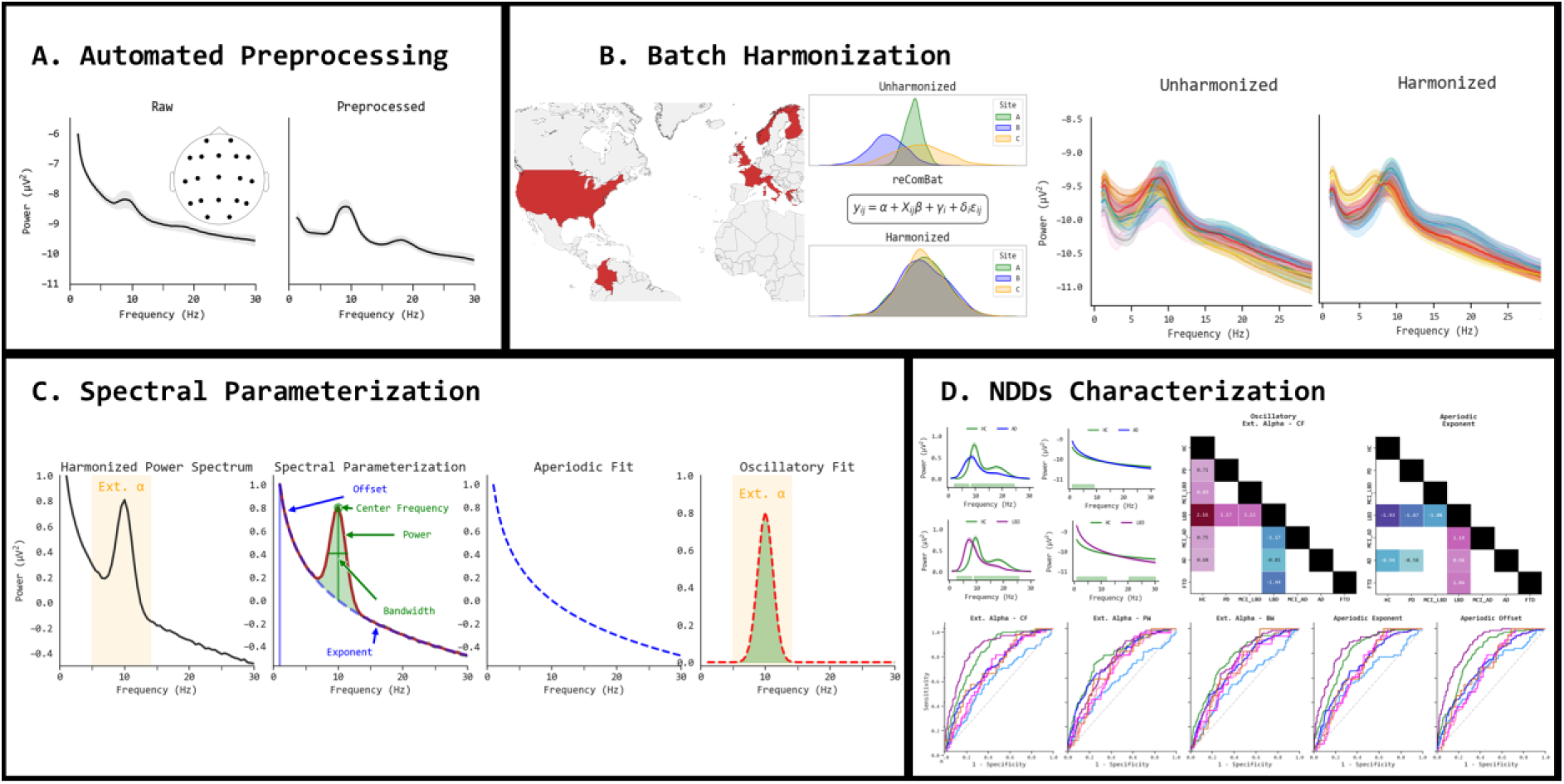
Graphical abstract of methods. **(A)** Resting-state EEG (rsEEG) data underwent an automated preprocessing regime comprising robust-average reference, bandpass filtering (1-45 Hz), wavelet-based independent component analysis for artifact rejection, downsampling to common electrode positions (19 channels) and frequency (128 Hz), bad epochs rejection based on statistical properties of the signal, and best epochs selection according to high power spectrum similarity. **(B)** Pooling multisite data (11 sites; 8 countries) introduces site-related variability (batch effects) in the distributions of rsEEG derivative features. Batch effects on the rsEEG power spectrum were assessed and subsequently mitigated using the reComBat algorithm (adjusting for age, sex and diagnosis-related variability). **(C)** On downstream analyses, the batch-harmonized rsEEG power spectrum was parameterized to separate oscillatory and aperiodic activity fit vectors, as well as spectral parameters (center frequency, power, bandwidth, aperiodic exponent, and aperiodic offset). **(D)** Characterization of oscillatory and aperiodic rsEEG activity fit vectors in neurodegenerative diseases (NDDs) was assessed with functional data analysis and mass-univariate statistics. Differences in the derivative spectral parameters across NDDs were evaluated using bootstrapped pairwise comparisons. Multinomial logistic regression models examined the separability across NDDs based on spectral parameters. **HC:** Healthy Controls; **MCI-AD:** Mild Cognitive Impairment – AD; **AD:** Alzheimer’s Disease; **PD:** Parkinson’s Disease; **MCI-LBD:** Mild Cognitive Impairment – Lewy Body Diseases; **LBD:** Lewy Body Dementias; **FTD:** Frontotemporal Dementia.

This manuscript adheres to the STROBE statement guidelines to provide clear reporting (Vandenbroucke et al., 2007). The codes used for analysis and figures will be made available on GitHub upon peer-reviewed publication of the manuscript.

## Results

### Demographic characteristics of the sample

In the pooled sample (n = 639 subjects in 11 sites), a total of 321 females (50.23%) and 318 males (49.77%) subjects were included. Age ranged from 44 to 89 years (Mean = 70.13, SD = 9.72, Median = 71, IQR = 64 – 77). Although age was comparable among males and females (Welch’s F = 0.11, p = 0.75), statistically significant differences were observed across sites (Welch’s F = 44.57, df = 10, p-uncorrected < 0.001, partial eta-squared = 0.40), with younger subjects predominantly in the California and Oslo datasets (Games-Howell test p < 0.05). Similarly, statistically significant age differences across NDDs were observed (Welch’s F = 38.39, df = 6, p-uncorrected < 0.05, partial eta-squared = 0.25), with significantly younger individuals, particularly in the HC, PD, and MCI-LBD subgroups (Games-Howell test p < 0.05). See Figure 2A and Supplementary Tables 4 – 6. On the other hand, a statistically significant cross-site difference in the proportion of female and male participants was observed (Chi-squared = 38.24, p < 0.001, df = 10). Similarly, sex proportion significantly varied across NDDs (Chi-squared = 28.95, p < 0.001, df = 6). Absolute and relative frequencies of sex by site and group are presented in Figures 2A and 2B.

**Figure 2.**
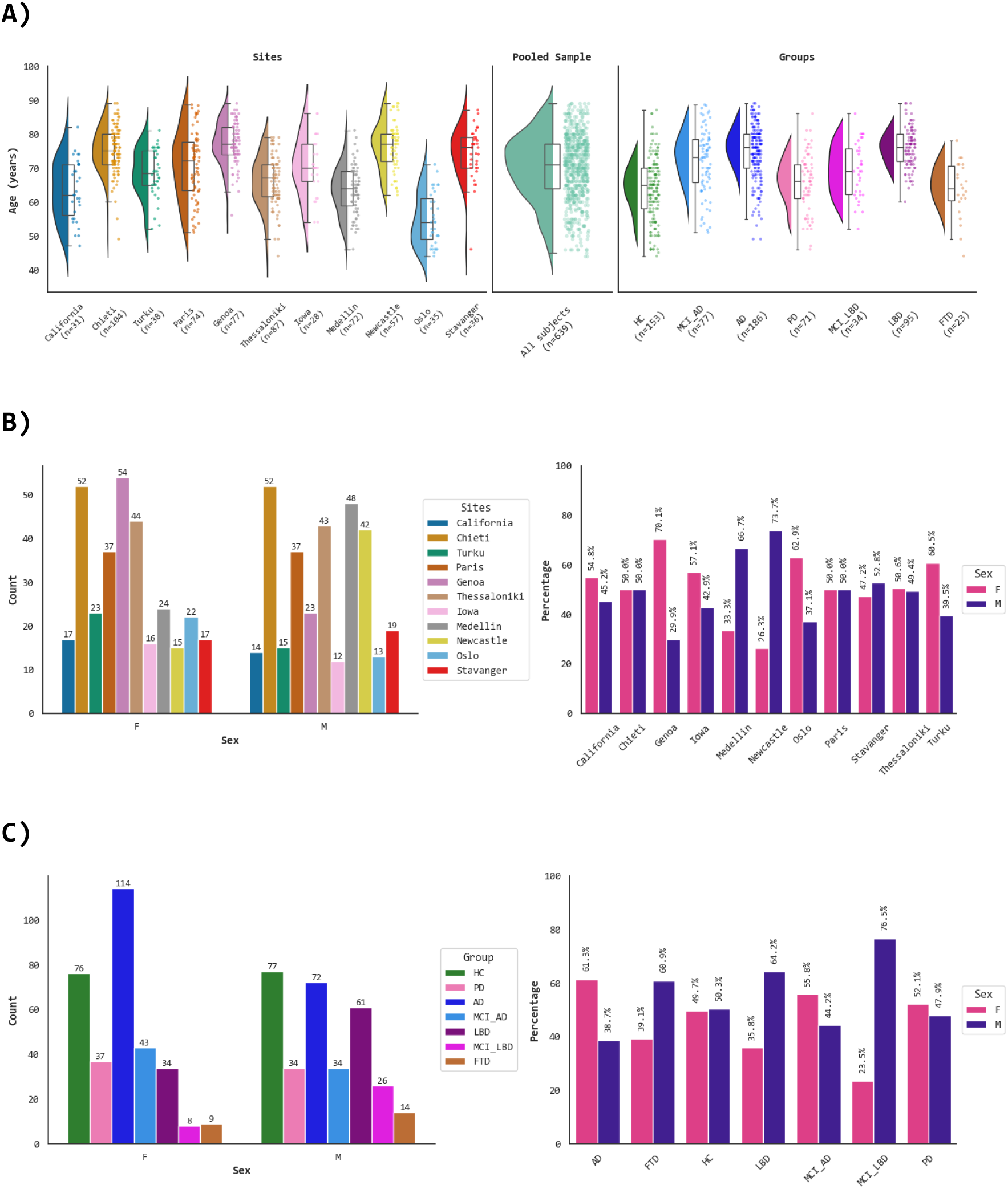
Demographical characteristics of the pooled sample (n = 639). **A)** From left to right, raincloud plots depict the age distributions by site, in the pooled sample, and by diagnosis groups. Bar plots show the absolute and relative frequency of female and male individuals across sites **(B)**, and diagnosis **(C)**. **F:** Female. **M:** Male. **AD:** Alzheimer’s Disease; **MCI-AD:** Mild Cognitive Impairment – AD; **PD:** Parkinson’s Disease; **MCI-LBD:** Mild Cognitive Impairment – Lewy Body Diseases (comprising MCI in PD and MCI with reported Lewy Body pathology); **LBD:** Lewy Body Dementias; **FTD:** Frontotemporal Dementia; **HC:** Healthy Controls.

### Batch-Harmonization of rsEEG Power Spectrum

Qualitative visualizations of batch effects in the posterior rsEEG power spectrum are presented in Figure 3 and Supplementary Figure 4. Univariate plots show the power spectrum distributions before and after harmonization. The distributions of batch-harmonized data were centered across sites, reducing the dispersion in some datasets (e.g., the Turku dataset), see Supplementary Figure 4. Bivariate plots suggested potential batch effects in the power spectrum with prominent variations in frequencies lower than 10 Hz. After batch-harmonization, the power spectrum vectors were better aligned across sites. Besides, PCA visualizations suggested less dispersed site centroids. Pairwise and cross-site average distances quantified on PCA components were lower in the batch-harmonized data; see Figure 3.

**Figure 3.**
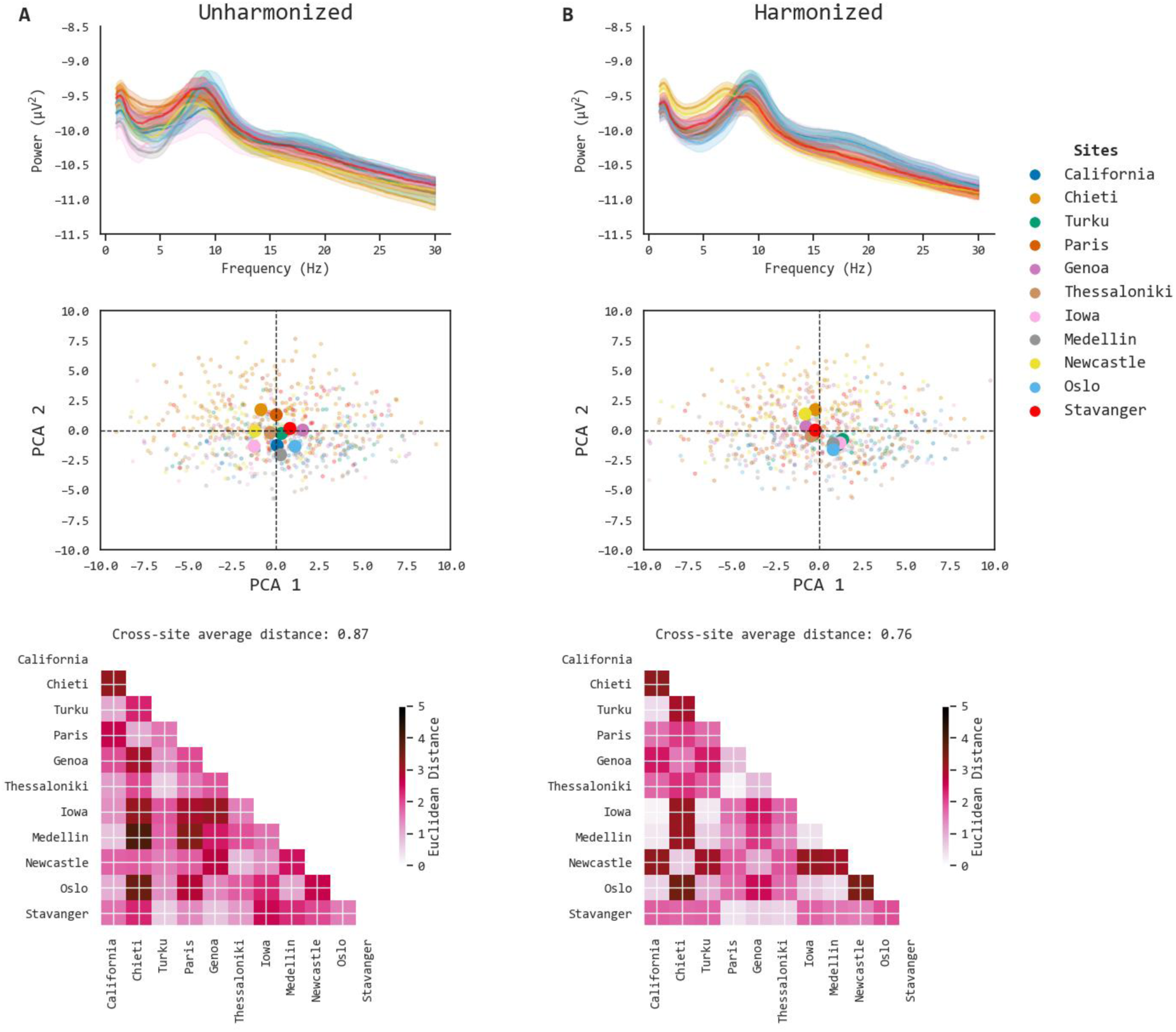
Inspection of batch and harmonization effects. The left column shows the unharmonized data (A), while the right side depicts the batch-harmonized data (B). The top row shows bivariate plots of the power spectrum across sites. Color lines illustrate the mean power spectrum in each site and its standard error with a 95 % confidence interval (dashed region). The second row shows Principal Component Analysis (PCA) plots by site. Each subject is represented in a single point, color-coded by the site. Bigger bold dots represent the centroid of each site. The bottom row presents the pairwise distance matrixes (Euclidean distances) between site centroids as a descriptor of batch effects. The cross-site average distance is computed from each matrix representing the overall batch effects in the unharmonized and harmonized data.

In line with qualitative visualizations, functional and mass-univariate ANOVAs supported the hypothesis of pre-existing statistically significant batch effects that were reduced after batch harmonization. Cross-site differences were observed across the whole bandwidth in the unharmonized data, with greater F-values in frequency bands lower than alpha. The batch-harmonized power spectrum exhibited greater differences in the theta band, followed by beta and alpha bands, see Figure 4.

**Figure 4.**
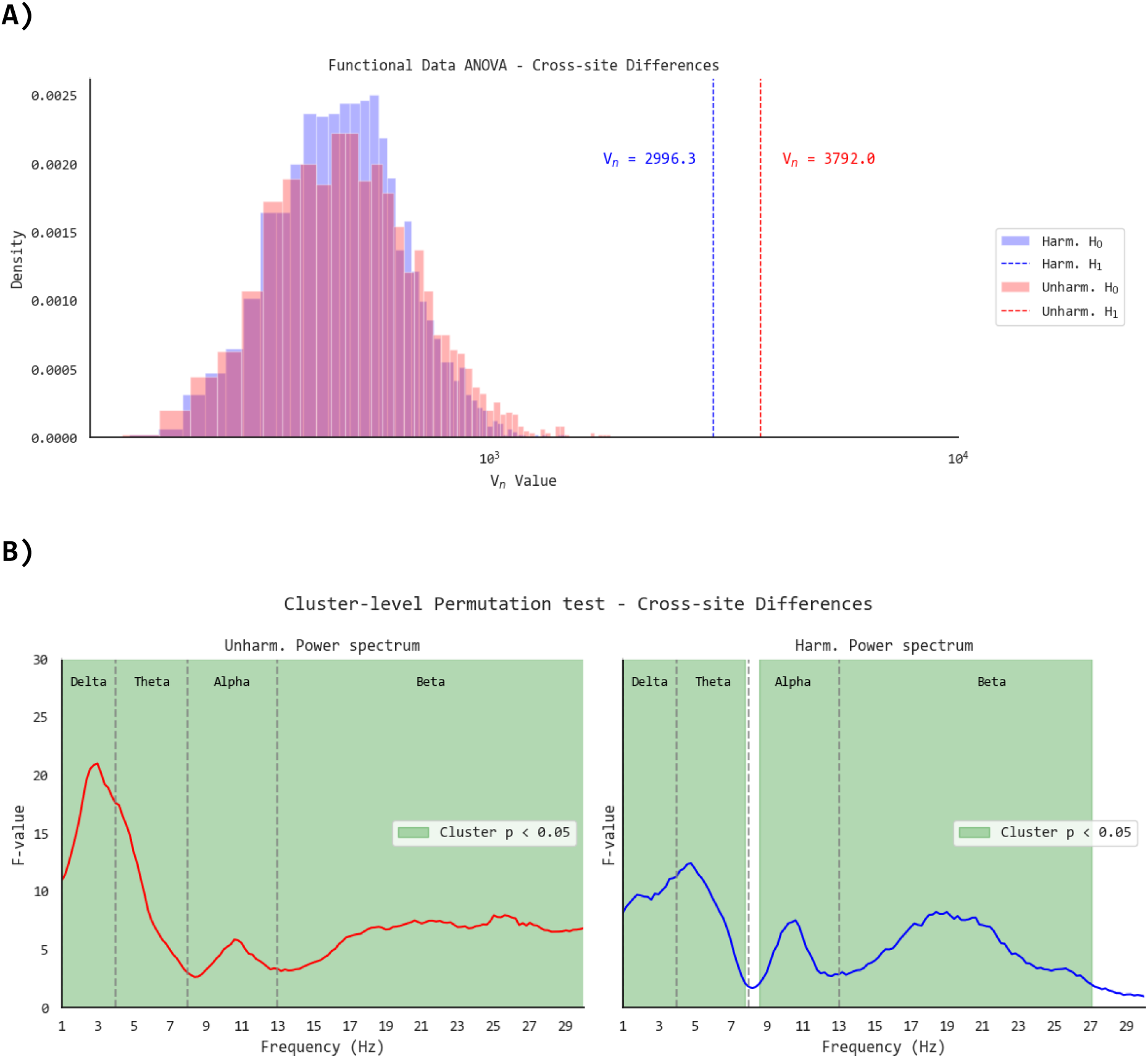
Statistical testing across batches. **(A)** Functional Data – One-way ANOVA with “site” as the independent variable and power spectrum (modeled through cubic B-spline basis, number of basis functions = 13) as the dependent variable. Histograms show a simulated distribution of the null hypothesis (i.e., no batch effects) in the unharmonized (Unharm. H_0_, red) and the harmonized power spectrum (Harm. H_0_, blue). The V_n_ statistic is an asymptotic version of the classical ANOVA F-value, representing the variability between batches. Dashed vertical lines depict the estimated V_n_ statistic testing the hypothesis of batch effects (H_1_) before (red) and after harmonization (blue); higher V_n_ values represent larger batch effects. **(B)** Mass-univariate permutation test for batch effects on power spectrum (2000 permutations, clustered on frequencies). F-values represent batch effects before (red) and after harmonization (blue); higher F-values represent larger batch effects (F-values Unharmonized – H_1_, min = 2.62 max = 21.02; F-values Harmonized – H_1_, min = 0.93 max=12.42). Statistically significant differences at the cluster level are highlighted in green.

In addition, functional and mass-univariate ANOVAs consistently indicated that cross-diagnosis differences were greater in the batch-harmonized power spectrum. Of note, the unharmonized power spectrum showed the most prominent cross-diagnosis differences in the delta, theta, and beta bands, but did not show significant results in the alpha band. By contrast, the batch-harmonized power spectrum revealed statistically significant cross-diagnosis differences across most of the frequency range, see Figure 5.

**Figure 5.**
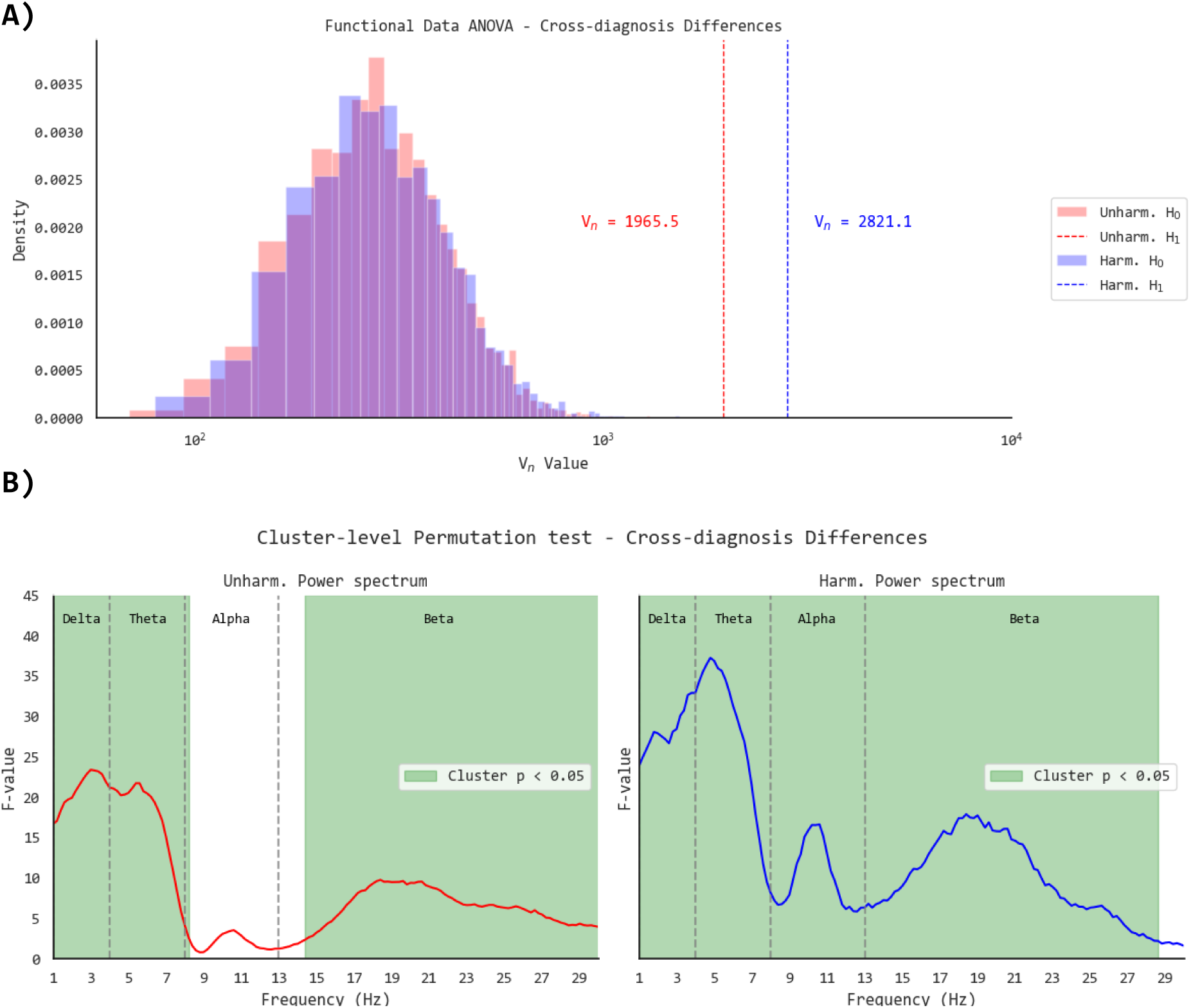
Statistical testing across diagnosis subgroups. **(A)** Functional Data – One-way ANOVA with “site” as the independent variable and power spectrum (modeled through cubic B-spline basis, number of basis functions = 13) as the dependent variable. Histograms show a simulated distribution of the null hypothesis (i.e., no batch effects) in the unharmonized (Unharm. H_0_, red) and the harmonized power spectrum (Harm. H_0_, blue). The V_n_ statistic is an asymptotic version of the classical ANOVA F-value, representing the variability between batches. Dashed vertical lines depict the estimated V_n_ statistic testing the hypothesis of batch effects (H_1_) before (red) and after harmonization (blue); higher V_n_ values represent larger batch effects. **(B)** Mass-univariate permutation test for batch effects on power spectrum (2000 permutations, clustered on frequencies). F-values represent batch effects before (red) and after harmonization (blue); higher F-values represent larger batch effects (F-values Unharmonized – H_1_, min = 0.79 max = 23.38; F-values Harmonized – H_1_, min = 1.55 max=37.23). Statistically significant differences at the cluster level are highlighted in green.

Pairwise differences across NDDs in the unharmonized and batch-harmonized power spectrum are depicted in Supplementary Figure 5. Altogether, qualitative visualizations and statistical testing supported the hypothesis of reduced batch effects after harmonization of the rsEEG power spectrum.

### Oscillatory and Aperiodic Activity across NDDs

The parameterization of the batch-harmonized power spectrum exhibited good fitting (R-squared greater or equal to 0.8) in most subjects (97.97%; n = 626), as shown in Supplementary Figures 6 and 7.

Pairwise differences across NDDs in the derivative fitting vectors of oscillatory and aperiodic activity are presented in Figure 6. Statistically significant clusters of reduced oscillatory alpha-band power (8-13 Hz) were observed in AD, compared to most NDDs except for the FTD group. We found a significant shifting of the oscillatory activity peak to frequencies lower than alpha in the LBD group, compared to other NDDs. Oscillatory activity between PD, MCI-LBD, and MCI-AD was not statistically different, see Figure 6A. The lowest peak frequency was observed in the LBD group (7.4 Hz), while the AD group exhibited the lowest peak power values, see Supplementary Figure 8. On the other hand, significant clusters of greater aperiodic activity were found in frequencies lower than alpha in LBD, compared to other diagnosis groups, see Figure 6B.

**Figure 6.**
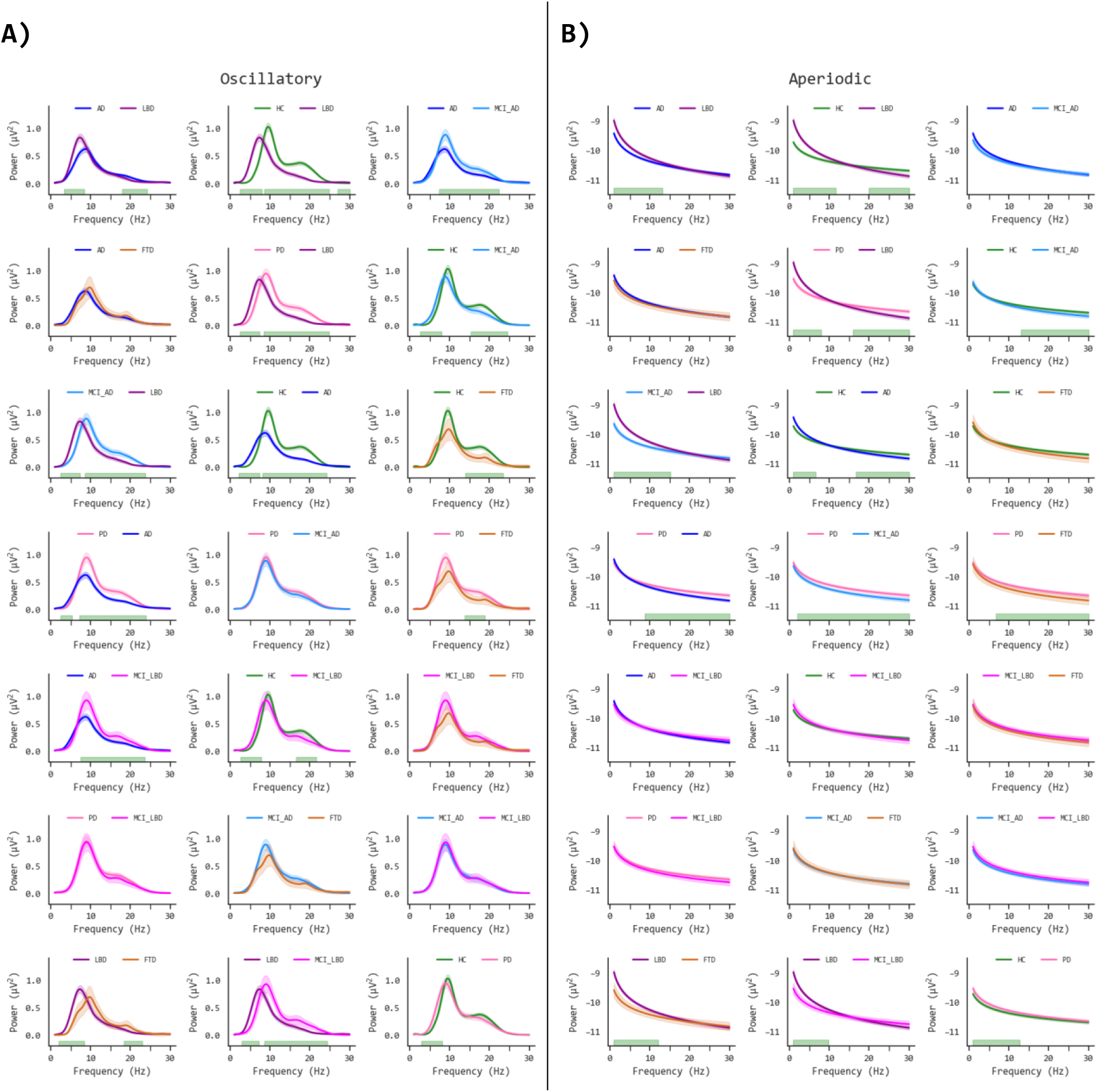
Pairwise comparisons of parameterized oscillatory aperiodic activity across neurodegenerative diseases (NDDs). **(A)** Mean oscillatory activity fits (lines) and 95 % standard error (shaded area); green regions on the x-axis bottom represent significant p values ( p < 0.05) on mass univariate permutation F-value tests (1000 permutations) clustered on frequencies. **(B)** Mean aperiodic activity fits (lines) and 95 % standard error (shaded area); green regions on the x-axis bottom represent significant clusters (p < 0.05). **HC:** Healthy Controls; **FTD:** Frontotemporal Dementia; **AD:** Alzheimer’s Disease; **PD:** Parkinson’s Disease; **LBD:** Lewy Body Dementia (comprising dementia in PD and Dementia with Lewy Bodies – DLB); **MCI-LBD:** Mild Cognitive Impairment in Lewy Body Dementia (comprising MCI in PD and MCI with reported Lewy Body pathology); **MCI-AD:** Mild Cognitive Impairment with reported AD pathology (or without Lewy Bodies).

Complementarily, we assessed pairwise comparisons on spectral parameters estimated from the derivative fitting vectors of oscillatory and aperiodic activity, see Figure 7.

**Figure 7.**
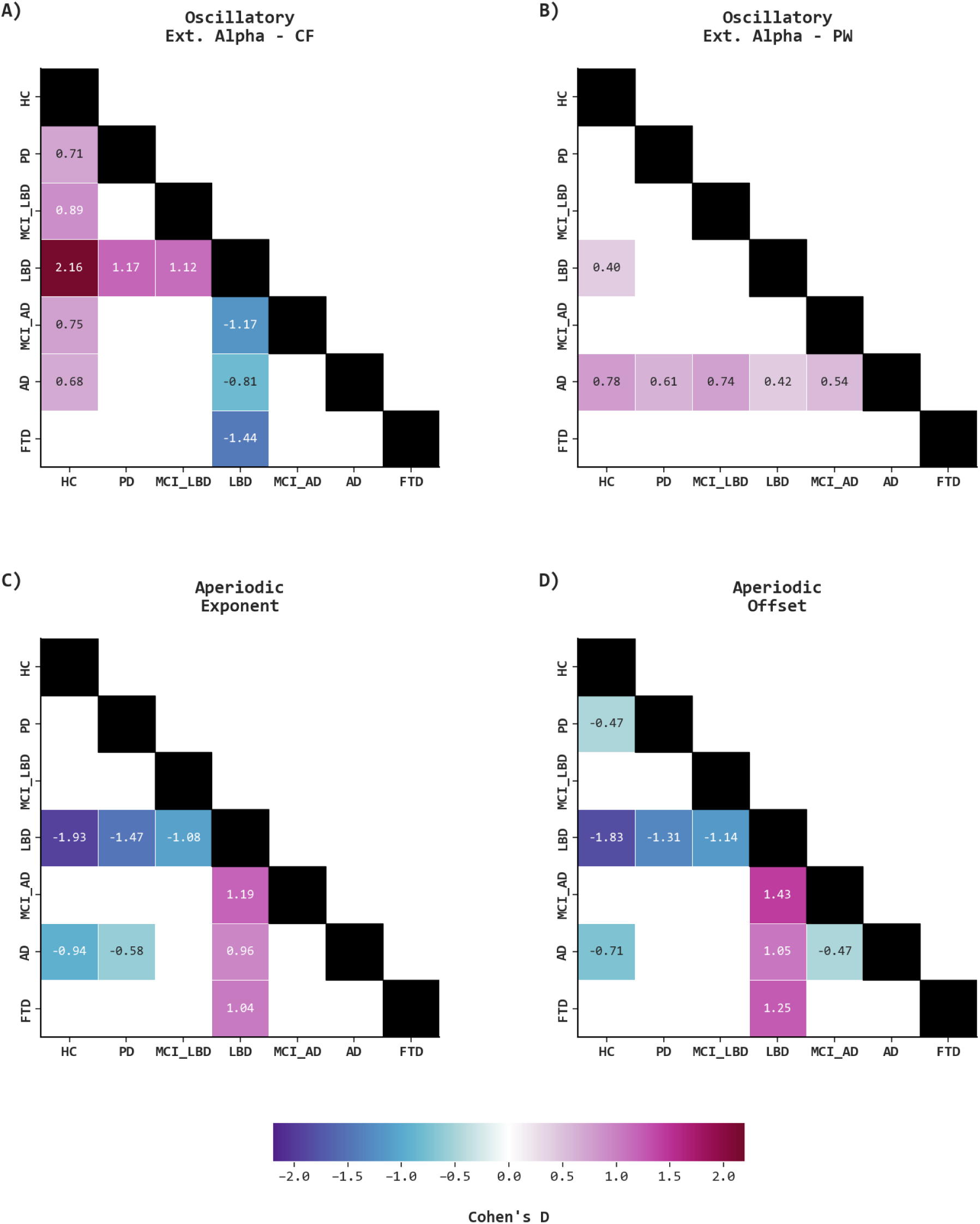
Differences in Spectral Parameters across neurodegenerative diseases (NDDs). Subplots present matrixes with the effect size (Cohen’s D) of pairwise differences for each derivative oscillatory (top) and aperiodic (bottom) spectral parameter: (A) Oscillatory Extended Alpha Center Frequency, (B) Oscillatory Extended Alpha Power, (C) Aperiodic Exponent, and (D) Aperiodic Offset. Effect sizes were calculated from parametric t-tests with bootstrapped confidence intervals (1000 iterations). P-values were corrected for multiple tests using the Benjamini-Yekutieli procedure. Non-statistically significant findings (i.e., corrected p-value greater or equal to 0.05) were masked in white. **HC:** Healthy Controls; **FTD:** Frontotemporal Dementia; **AD:** Alzheimer’s Disease; **PD:** Parkinson’s Disease; **LBD:** Lewy Body Dementia (comprising dementia in PD and Dementia with Lewy Bodies – DLB); **MCI-LBD:** Mild Cognitive Impairment in Lewy Body Dementia (comprising MCI in PD and MCI with reported Lewy Body pathology); **MCI-AD:** Mild Cognitive Impairment with reported AD pathology (or without Lewy Bodies); **Ext. Alpha:** Extended Alpha band; **CF:** Center Frequency; **PW:** Power.

Oscillatory extended alpha CF showed significant differences between LBDs and other groups with large effect sizes. Similarly, extended alpha CF was significantly different in HCs vs all NDDs comparisons (except for FTD) exhibiting moderate to large effect sizes, see Figure 7A. Besides, significantly lower extended alpha PW was observed when comparing AD vs other groups with moderate effect sizes, see Figure 7B. Extended alpha BW did not yield statistically significant findings. Further, significant results on aperiodic parameters supported consistent differences in LBD vs. other groups with large effect sizes on the aperiodic exponent (Figure 7C) and aperiodic offset (Figure 7D). Forest plots of bootstrapped pairwise differences in the derivative oscillatory and aperiodic parameters are presented in Supplementary Figures 9 and 10.

Finally, Receiver Operating Characteristics (ROC) curves assessing the discriminatory ability of derivative spectral parameters in the separation of NDDs are presented in Figure 8. Unadjusted multinomial logistic regression models (each spectral parameter as a single predictor of diagnosis) showed good discrimination particularly in the LBD individuals when compared to other groups. Thus, in the LBD group, the highest area under the ROC curve (AUC) was observed in the Aperiodic Offset (Cutoff-Youden Index = –9.49; AUC = 0.83; Sensitivity = 0.82; Specificity = 0.69; Positive Predictive Value – PPV = 0.32; Negative Predictive Value – NVP = 0.96), followed by the Aperiodic Exponent (Cutoff-Youden Index = 1.07; AUC = 0.82; Sensitivity = 0.75; Specificity = 0.76; PPV = 0.36; NVP = 0.95), and the Extended Alpha CF (Cutoff-Youden Index = 6.03; AUC = 0.81; Sensitivity = 0.90; Specificity = 0.63; PPV = 0.30; NVP = 0.97). Age– and sex-adjusted models yielded consistent findings with similar or marginally increased AUCs.

**Figure 8.**
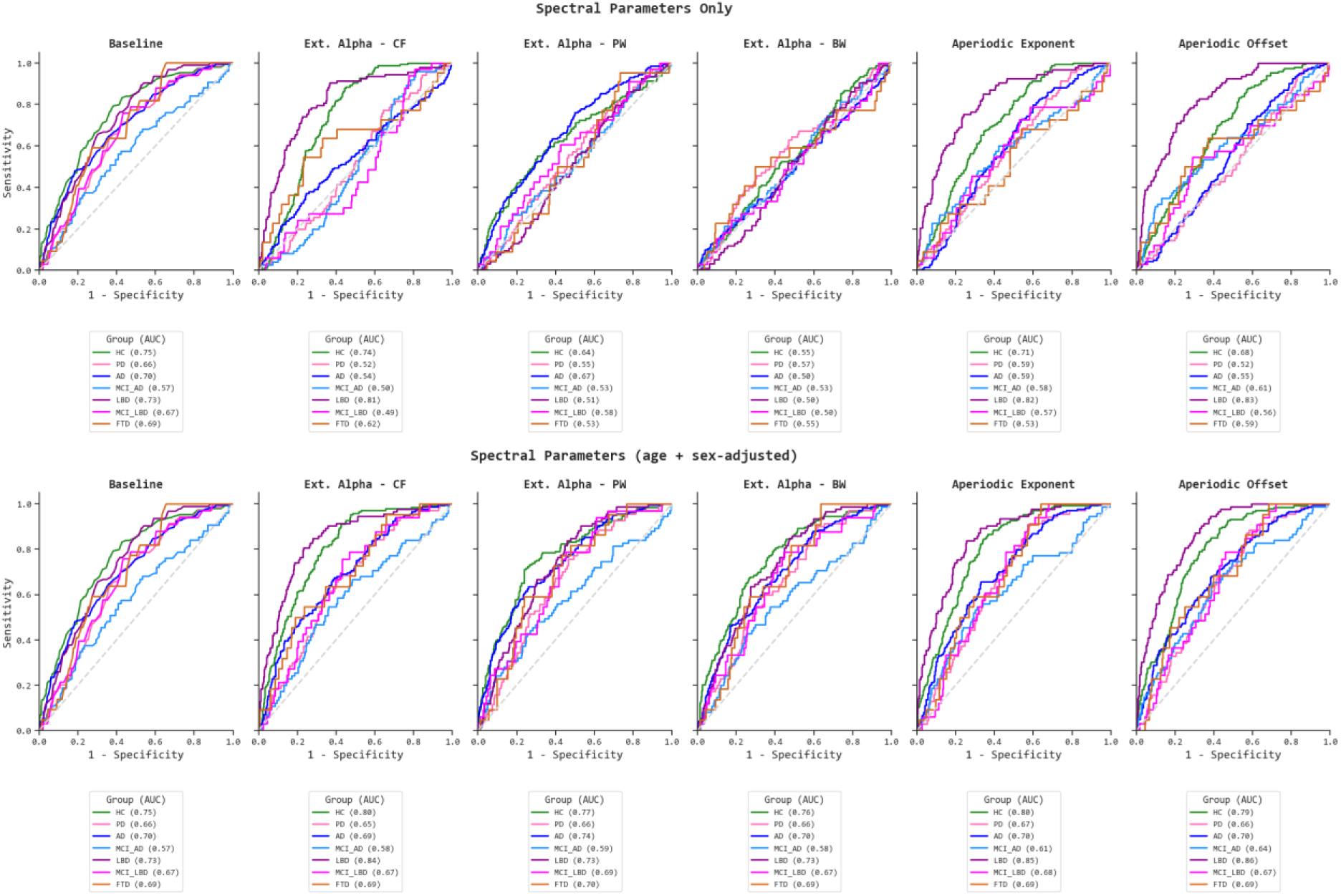
Associations Between Spectral Parameters and Neurodegenerative Diseases (NDDs) Diagnosis. Unadjusted (top) and age + sex-adjusted results (bottom) from multinomial logistic regressions showing the Receiver Operating Characteristic (ROC) curves for each Spectral Parameter as a predictor of the NDDs groups. A baseline model with age and sex as predictors of diagnosis was also fitted. **HC:** Healthy Controls; **FTD:** Frontotemporal Dementia; **AD:** Alzheimer’s Disease; **PD:** Parkinson’s Disease; **LBD:** Lewy Body Dementia (comprising dementia in PD and Dementia with Lewy Bodies – DLB); **MCI-LBD:** Mild Cognitive Impairment in Lewy Body Dementia (comprising MCI in PD and MCI with reported Lewy Body pathology); **MCI-AD:** Mild Cognitive Impairment with reported AD pathology (or without Lewy Bodies); **AUC:** Area Under the ROC curve; **Ext. Alpha:** Extended Alpha band; **CF:** Center Frequency; **PW:** Power; **BW:** Bandwidth.

Based on the AUC ROC, the highest discrimination of LBD individuals was achieved by the Aperiodic Offset (Cutoff-Youden Index = –9.41; AUC = 0.86; Sensitivity = 0.87; Specificity = 0.70; PPV = 0.34; NVP = 0.97), followed by the Aperiodic Exponent (Cutoff-Youden Index = 0.90; AUC = 0.85; Sensitivity = 0.84; Specificity = 0.74; PPV = 0.36; NVP = 0.96), and the Extended Alpha CF (Cutoff-Youden Index = 7.04; AUC = 0.84; Sensitivity = 0.81; Specificity = 0.77; PPV = 0.38; NVP = 0.96). Similarly, age– and sex-adjusted models achieved good discrimination of HC individuals when using the Extended alpha CF (Cutoff-Youden Index = 8.82; AUC = 0.80; Sensitivity = 0.91; Specificity = 0.57; PPV = 0.40; NVP = 0.95) or the Aperiodic Exponent (Cutoff-Youden Index = 0.58; AUC = 0.80; Sensitivity = 0.85; Specificity = 0.66; PPV = 0.44; NVP = 0.93) as predictors of diagnosis. No other predictors exhibited good or high performance (AUC ROC greater or equal to 0.80) in the discrimination of NDDs. A comprehensive summary of performance metrics obtained from multinomial logistic regression models is presented in Supplementary Figure 11.

## Discussion

In this multicentric study, we characterized spectral patterns across multiple clinical phenotypes of NDDs by isolating the oscillatory and aperiodic activity from the rsEEG power spectrum. Besides, we assessed and controlled for potential site-related batch effects that could confound the downstream analysis of rsEEG derivative features. We found that batch-harmonization can effectively mitigate site-related differences on the rsEEG power spectrum and preserve diagnosis-related differences (with an increased effect size). Notably, spectral parameterization exhibited a distinctive pattern in the LBD group, characterized by the prominent slowing (< 7.3 Hz) of the extended alpha center frequency in the posterior channels with greater aperiodic activity (steeper exponents and higher offsets). Complementarily, the lower posterior extended alpha power was a signature of AD (with smaller effects on center frequency and aperiodic parameters). Our findings support the external validity of previous smaller studies reporting oscillatory alpha power differences in AD (Kopčanová et al., 2024; Wang et al., 2024), and oscillatory alpha slowing with abnormal aperiodic activity in alpha-synuclein-related disorders (including PD and LBD) (Burelo et al., 2024; McKeown et al., 2023; Rosenblum et al., 2023). Given the relevance of abnormal posterior alpha rhythms in NDDs, spectral parameterization is crucial to measure oscillatory activity (without conflation of the underlying aperiodic activity) and to explore candidate patterns for differential diagnosis across clinical phenotypes. This expands the current evidence mainly focused on AD and PD populations, as highlighted by recent systematic reviews (Donoghue, 2024; Fernández-Rubio et al., 2024). By pooling multisite datasets while controlling for batch effects, we provide a robust pipeline to tackle replicability issues arising from small sample sizes (Button et al., 2013) typically observed in rsEEG studies assessing pathological aging (Fernández-Rubio et al., 2024; Newson and Thiagarajan, 2019).

### Batch-Harmonization of rsEEG Power Spectrum

Convergent evidence obtained from multisite datasets has demonstrated batch effects on the rsEEG time series (Bigdely-Shamlo et al., 2020), and derived features, including spectral parameters (Jaramillo-Jimenez et al., 2024), functional connectivity (Moguilner et al., 2022; Prado et al., 2022), power spectra and Riemannian geometry embeddings computed from channel covariance matrixes (Li et al., 2022; Mellot et al., 2024, 2023). Consistent with these reports, our results on unharmonized rsEEG power spectra revealed significant batch effects affecting the 1 – 30 Hz frequency range (with greater effect sizes in the delta band compared to the theta, alpha, and beta bands). Batch-harmonization reduced the overall magnitude of batch effects (see Figure 4) achieving better cross-site alignment of the power spectrum. Moreover, batch harmonization enhanced diagnosis-related differences in the alpha band, while preserving preexisting differences in the delta, theta, and beta bands (see Figure 5).

Insights from multicentric neuroimaging collaborations recommend modeling site-specific parameters to statistically correct potential batch effects (Hu et al., 2023). Among the most widely used methods for statistical batch effects correction, ComBat-derived algorithms have gained traction as reliable and straightforward tools for multicentric harmonization, suitable for inferential statistics and machine learning predictive modeling (Bell et al., 2022; Da-ano et al., 2020; Horng et al., 2022a; Hu et al., 2023; Marzi et al., 2024). Inspired by the success of ComBat-based algorithms for harmonization of batch effects (related to site, scanner, or radiotracer) across multiple features and data modalities — and their scarce application in electrophysiological data (Li et al., 2022) — we benchmarked these harmonization methods in rsEEG spectral parameters CF, PW, BW, aperiodic exponent, and offset (Jaramillo-Jimenez et al., 2024). However, our previous study assessed rsEEG age-related changes in healthy subjects only, not including ComBat-based algorithms capable of handling a singularity of the design matrix with biological covariates (e.g., adjusting for site and diagnosis effects when all subjects at a given site share the same diagnosis). Following this rationale, when site and diagnosis represent the same population, batch effects become undistinguishable from biological covariate effects and cannot be removed with traditional ComBat implementations. Although not frequently discussed, design matrix singularity issues could be expected when repurposing or pooling several retrospective datasets. To overcome this, the reComBat model estimates site-related parameters with regularization techniques that allow the resolution of singular design matrixes (Adamer et al., 2022). Interestingly, batch effects in the median posterior power spectra were not as prominent as those previously observed on rsEEG spectral parameters (Jaramillo-Jimenez et al., 2024). The latter might be explained as the distribution of the power spectrum vector tends to normality after log-transformation, and it has smaller scale variations compared to spectral parameters such as CF (ranging from 7.3 – 9.5 Hz), PW (0.8 – 1.2 microVolts), or offset (–9.7 – –8.9 microVolts), which might result in larger batch-effects more evident on qualitative visualizations. Although the present results support the hypothesis of reduced batch-related differences on the rsEEG power spectrum after harmonization, we recognize that further research is needed to shed light on knowledge gaps including the effect of unbalanced covariates across batches, transfer-learning approaches for unseen batches, preserving dependencies on data structure (e.g., covariance, shape and spatial patterns), or handling some limitations of ComBat based algorithms such as outliers, extreme values, multimodal distributions, among others (Cetin-Karayumak et al., 2020; Han et al., 2023).

### Slow oscillatory alpha frequency and increased aperiodic activity in LBD

Seminal publications have shown consistent rsEEG spectral patterns in LBD (Bonanni et al., 2016, 2008; Chatzikonstantinou et al., 2021; van der Zande et al., 2018), characterized by a reduced posterior dominant peak frequency (< 8 Hz) with or without increased dominant frequency variability. These findings were incorporated as part of the current supportive diagnostic criteria for dementia and prodromal stages of LBD (McKeith et al., 2020, 2017). Although most studies converged on LBD-related abnormalities in posterior oscillatory activity, batch effects were typically overlooked and not addressed in prior research (Watanabe et al., 2024). Our findings in the LBD group were compatible between the unharmonized and batch-harmonized power spectrum, reflecting significant pairwise differences in the extended alpha frequency band. Though oscillatory findings in alpha-synucleinopathies could be undermined by confounding aperiodic activity, typically not parameterized in LBD clinical research (Donoghue, 2024; Donoghue et al., 2020), few recent publications have performed rsEEG spectral parameterization on small sample size data, showing steeper aperiodic activity in PD, and even greater in LBD (Burelo et al., 2024; McKeown et al., 2023; Rosenblum et al., 2023). Along with the high aperiodic activity of alpha-synucleinopathies documented in preliminary reports, a prominently low oscillatory center frequency (in the 4 – 15 Hz range) prevails as a robust hallmark of LBD when compared to cognitively normal PD, MCI, and AD groups (Burelo et al., 2024; Rosenblum et al., 2023). Consistently, our results elucidated large pairwise differences in LBD, with the largest effect size observed for reduced oscillatory extended alpha CF, followed by aperiodic exponent and aperiodic offset. Although aperiodic parameters were similar among MCI-LBD and PD groups, MCI-LBD vs. LBD, PD vs. LBD, and HC vs. LBD comparisons yielded a significant pattern of low aperiodic activity in HC, moderate in MCI-LBD and PD, and high in LBD.

Current models propose a neurophysiological basis for aperiodic activity by accounting for the interaction of synaptic kinetics, excitatory/inhibitory balance, aperiodic network dynamics, and non-rhythmic neural activity (Brake et al., 2024). Albeit augmented (steeper) aperiodic activity in LBD has been hypothesized as a surrogate biomarker of increased inhibition, consensus on these conclusions is not sustained, and future research with more control of experimental designs is required to clarify these relevant aspects. For a detailed discussion on aperiodic activity in clinical research, the reader is referred to a recent systematic review (Donoghue, 2024).

### Low oscillatory alpha PW in AD

Reduced alpha power and peak frequency along with increasing delta and theta power have been reported as consistent signatures of AD in clinical (Babiloni et al., 2011; Brueggen et al., 2017; Modir et al., 2023; Moretti, 2015; Moretti et al., 2004; Triggiani et al., 2017) and simulation studies (Alexandersen et al., 2023). Neurophysiological data exhibiting synergistic interactions with neuropathology (Gallego-Rudolf et al., 2024), and cerebrospinal fluid (CSF) biomarkers (Smailovic et al., 2018) of AD-related pathology have also contributed to expert recommendations supporting spectral analysis of the delta-theta and alpha band powers in combination with the alpha peak frequency as promising markers of AD and prodromal MCI-AD stages (Babiloni, 2022; Babiloni et al., 2021, 2020a). The validity of findings related to alpha oscillations has been repeatedly assessed via spectral parameterization of the power spectrum, often achieving improved associations between oscillatory activity and clinical features of interest (Donoghue, 2024; Kopčanová et al., 2024). As a case in point, a previous study found reduced oscillatory alpha PW with generalizable results in two small sample size cohorts (Cohort 1 n = 45, AD = 18; Cohort 2 n = 31, AD = 25), whereas aperiodic parameters were comparable among AD and HCs. Similarly, one previous investigation of the Thessaloniki dataset (included in our study), parameterized the rsEEG power spectrum in AD (n = 36) vs HC (n = 29), observing significantly lower oscillatory alpha PW in AD (with increased F-values after spectral parameterization). Nonetheless, these authors also reported increased posterior aperiodic exponent and offsets in AD (Wang et al., 2024). Our observations on reduced oscillatory alpha PW in AD (with significant moderate-to-large effect sizes across all NDDs) support earlier reports. Aperiodic findings were less concluding, supporting increased aperiodic exponent and offset in AD, compared to HC (Donoghue, 2024; Wang et al., 2024). Inconsistent aperiodic findings in AD have been attributed to i) potential sample heterogeneity in disease etiology and severity, and ii) a dynamic response of aperiodic parameters across the progression of the AD continuum not reflected in a gross clinical phenotype. Multimodal data integration could emerge as a promising strategy to fill current knowledge gaps on the association between clinical, biological, and neurophysiological trajectories of AD (Hebling Vieira et al., 2022; Prado et al., 2023).

## Limitations

Standardized experimental conditions are crucial to enhance comparability when pooling multisite datasets in clinical investigations. However, this was not possible as we were not directly involved in the design of all primary studies. In addition, the scarcity of longitudinal rsEEG data precluded us from the estimation of change trajectories, or test-retest reliability assessments. In line with this, clinical diagnosis of NDDs was performed by specialized clinician evaluations based on operationalized criteria, although similar, those criteria varied across sites, potentially resulting in increased clinical and biological heterogeneity. Sensitivity analyses using biological definitions of AD (Aisen et al., 2017), PD, and LBD (Simuni et al., 2024) could complement results derived from clinical phenotypes of neurodegeneration.

We selected methods accounting for robust estimators (based on iterative statistics), nonetheless, present results were conducted at the group level, requiring future dedicated studies to evaluate the performance of batch-harmonized spectral parameters for the individual-level classification of NDDs. Besides, the reader must consider that our scope was focused on NDDs rather than age-related changes in spectral parameters. The latter was comprehensively assessed by our group in a prior publication (Jaramillo-Jimenez et al., 2024). Beyond this, we did not estimate source-space reconstructions due to their associated computational demand, which complicates the local analysis of large-scale datasets. Even if sensor space features reflect mixed source activity across adjacent channels due to volume conduction, we supported our selection of posterior channels, as occipital dipoles are the most relevant generators of the posterior rsEEG power spectrum (Schaworonkow and Nikulin, 2022). Finally, extensive benchmarking of the reComBat model using synthetic data as ground truth fell outside the scope of our study but could be explored by researchers working with pooled multisite datasets (Marzi et al., 2024). Other factors potentially affecting the batch harmonization, include cross-batches sample size differences (Parekh et al., 2022), outlier effects (Han et al., 2023) and data leakage when using ComBat-derived methods without dedicated implementations for predictive machine learning models (Marzi et al., 2024).

## Conclusions

This study capitalizes on multicentric data accounting for diverse clinical phenotypes of NDDs. We present a detailed characterization of oscillatory and aperiodic activity while addressing limitations from preliminary publications, especially in LBD clinical research. Our observations support that batch effects in the rsEEG power spectrum can be mitigated with harmonization while preserving (and increasing) diagnosis-related differences. Oscillatory alpha PW reduction may better reflect AD abnormalities, whereas pronounced oscillatory CF slowing and greater aperiodic activity characterize the LBD group. We propose an adaptable open pipeline with a common preprocessing regime, batch harmonization, and spectral parameterization along with visualizations and statistical testing of batch, harmonization, and group-related effects. Further investigations can benefit from data pooling to build up larger datasets suitable for predictive modeling at the individual level.

## Data availability

Publicly available repositories host the open datasets (refer to the Participants section). Access to in-house clinical rsEEGs is restricted due to ethical considerations and can only be granted upon approval of a project proposal by the EDLB Steering Committee; for inquiries, please contact the corresponding author. All codes for data preprocessing and analysis will be made publicly available on a GitHub repository upon the peer review and acceptance of this pre-print.

## Supporting information

Supplementary Materias

## Acknowledgments

We extend our gratitude to all participants and patients who generously contributed their data to the dementia research community. Besides, we are particularly thankful to the investigators of the primary studies for embracing the principles of open science and making their datasets publicly accessible. Additionally, we acknowledge the invaluable support provided by institutions and organizations that have facilitated the research efforts of many authors of this paper. These include funding institutions (Helse Vest; Project number F-12155), research environments: Centre for Age-Related Medicine (SESAM) in Norway, EDLB consortium, Grupo de Neurociencias de Antioquia (GNA) and Grupo Neuropsicología y Conducta (GRUNECO), as well as research incubator programs such as the Semillero NeuroCo and Semillero SINAPSIS at the School of Medicine, University of Antioquia, Colombia.

## Author contributions

AJ-J (A, B, C, D, E, F, H, J, K, L); Y-JM-R (B, D, E, H, L); DAT-R (A, C, D, E, L); FL (F, G, I, L); DAg (F, G, I, L); JFO-G (E, F, G, H, I, L); CP (F, G, I, L); SG (E, F, G, L); MP (F, G, I, L); DAr (F, G, I, L); J-PT (F, G, I, L); TF (F, G, I, L); KB (F, G, I, L); DAa (F, G, I, L); LB (F, G, I, L). **A:** Conceptualization; **B:** Data Curation. **C:** Formal Analysis; **D:** Investigation; **E:** Methodology; **F:** Project Administration; **G:** Resources; **H:** Software; **I:** Supervision; **J:** Visualization; **K:** Writing – Original Draft; **L:** Writing – Review & Editing.

## Declaration of generative AI and AI-assisted technologies in the writing process

During the preparation of this work, ChatGPT 4 and ChatGPT 4o (by OpenAI) were used to improve readability. The authors reviewed and edited the resulting outputs and take full responsibility for the content of the publication.

