## Supplementary Materias for "Characterizing resting-state EEG oscillatory and aperiodic activity in neurodegenerative diseases: A multicentric study"

### Supplementary Materials

##### Supplementary Table 1. Data sources

| Dataset, Country | Cohort / Repository | Publication Digital Object Identifier (DOI) |
|---|---|---|
| **California, USA** | OpenNeuro | https://doi.org/10.18112/openneuro.ds002778.v1.0.5 |
| **Chieti, ITA** | EDLB | https://doi.org/10.3233/jad-160435 |
| **Turku, FIN** | OSF | https://osf.io/pehj9/ |
| **Paris, FRA** | EDLB | N.A. |
| **Genoa, ITA** | EDLB | https://doi.org/10.3233/jad-160435 |
| **Newcastle, GBR** | EDLB | https://doi.org/10.1186/s13195-020-00650-1 |
| **Thessaloniki, GRC** | OpenNeuro | https://doi.org/10.18112/openneuro.ds004504.v1.0.7 |
| **Iowa, USA** | Dropbox | https://doi.org/10.1016/j.parkreldis.2020.08.001 |
| **Medellin, COL** | N.A. | https://doi.org/10.1016/j.clinph.2021.01.001 |
| **Oslo, NOR** | OpenNeuro | https://doi.org/10.18112/openneuro.ds003775.v1.2.1 |
| **Stavanger 1, NOR** | DDI | N.A. |
| **Stavanger 2, NOR** | EDLB | N.A. |

**EDLB:** European Dementia with Lewy Bodies Consortium; **OSF:** Open Science Framework; **DDI:** Dementia Disease Initiation Study; **NA:** Not publicly available.

**Supplementary Table 2. Diagnostic criteria used in Independent Datasets**

| Dataset, Country | Sample size (n) | NDDs subgroups (n) | Diagnostic criteria |
|---|---|---|---|
| **California, USA** | 31 | HC (16), PD (15) | Specialist in movement disorders; operationalized criteria not reported |
| **Chieti, ITA** | 104 | AD (38), LBD (63), MCI-AD (1), PD (2) | At least two core features of DLB criteria (McKeith et al., 1996; Mc Keith et al., 2005); National Institute of Neurological Disorders and Stroke–Alzheimer Disease and Related Disorders (NINCDS-ADRDA) (McKhann et al., 1984); MCI-AD defined as subjects fulfilling Mild Cognitive Impairment criteria (Petersen et al., 1999) without evidence of any DLB core/suggestive symptoms (Mc Keith et al., 2005) and no criteria of vascular dementia (Roman et al., 1993); United Kingdom Brain Bank criteria (Gibb and Lees, 1988). |
| **Turku, FIN** | 38 | HC (19), PD (19) | United Kingdom Brain Bank criteria (Gibb and Lees, 1988) or Movement Disorder's Society (MDS) criteria (Postuma et al., 2015) |
| **Paris, FRA** | 74 | MCI-AD (58), MCI-LB (16) | Confirmed Alzheimer's disease pathology on CSF (A+T+), criteria (Jack et al. 2018); Research criteria for the diagnosis of prodromal dementia with Lewy bodies (McKeith et al., 2020); |
| **Genoa, ITA** | 77 | AD (77) | Diagnostic and Statistical Manual of Mental Disorders, fourth edition (DSM-IV-TR) and National Institute of Neurological Disorders and Stroke–Alzheimer Disease and Related Disorders (NINCDS-ADRDA) (McKhann et al., 1984) |
| **Newcastle, GBR** | 57 | AD (32), LBD (25) | Fourth consensus revised DLB criteria (McKeith et al., 2017), Clinical criteria for dementia associated with Parkinson's Disease (Emre et al., 2007), National Institute on Aging-Alzheimer's Association (NIA-AA) (McKhann et al., 2011) |
| **Thessaloniki, GRC** | 87 | HC (28), AD (36), FTD (23) | Specialist team in neurology; operationalized criteria not reported |
| **Iowa, USA** | 28 | HC (14), MCI-PD (2), PD (12) | United Kingdom Brain Bank criteria (Gibb and Lees, 1988), Movement Disorder's Society Task Force on mild cognitive impairment in Parkinson's disease level 1 (Litvan et al., 2011) |
| **Medellin, COL** | 72 | HC (36), PD (22), MCI-PD (14) | Movement Disorder's Society (MDS) criteria (Postuma et al., 2015), MDS Task Force on mild cognitive impairment in Parkinson's disease level 1 (Litvan et al., 2011) |
| **Oslo, NOR** | 35 | HC (35) | N.A. |
| **Stavanger 1, NOR** | 23 | HC (5), MCI-AD (18) | National Institute on Aging-Alzheimer's Association (NIA-AA) for MCI due to AD with intermediate/high likelihood (Albert et al., 2011) |
| **Stavanger 2, NOR** | 13 | AD (3), LBD (7), MCI-LB (2), PD (1) | National Institute on Aging-Alzheimer's Association (NIA-AA) (McKhann et al., 2011); Fourth Consensus revised DLB criteria (McKeith et al., 2017); Clinical criteria for dementia associated with Parkinson's Disease (Emre et al., 2007); Research criteria for the diagnosis of prodromal dementia with Lewy bodies (McKeith et al., 2020); Movement Disorder's Society (MDS) Parkinson's Disease criteria (Postuma et al., 2015) |

**HC:** Healthy Controls; **FTD:** Frontotemporal Dementia; **AD:** Alzheimer's Disease; **PD:** Parkinson's Disease; **DLB:** Dementia with Lewy Bodies. **LBD:** Lewy Body Dementias (comprising dementia in PD and DLB); **MCI-LB:** Mild Cognitive Impairment with Lewy Bodies; **MCI-PD:** Mild Cognitive Impairment in PD; **MCI-AD:** Mild Cognitive Impairment with AD pathology (or without Lewy Bodies).

**Supplementary Table 3. RsEEG acquisition parameters used in Independent Datasets**

| Dataset, Country | Cohort / Repository | # Channels | Sampling Frequency (Hz) | Amplifier/Headset | Minimum # of clean epochs |
|---|---|---|---|---|---|
| **California, USA** | OpenNeuro | 32 | 512 | BioSemi ActiveTwo system | 30 |
| **Chieti, ITA** | EDLB | 21 | 128 | NEUROSCAN SynAmps | 71 |
| **Turku, FIN** | OSF | 64 | 500 | NeurOne Tesla | 21 |
| **Paris, FRA** | EDLB | 19 | 256 | MICROMED SYSTEM PLUS | 20[*] (3) |
| **Genoa, ITA** | EDLB | 19 | 128 - 256 | EBNeuro BE Plus LTM | 20 [*] (3) |
| **Newcastle, GBR** | EDLB | 128 | 1024 | ANT Neuro | 23 |
| **Thessaloniki, GRC** | OpenNeuro | 19 | 500 | Nihon Kohden EEG 2100 | 48 |
| **Iowa, USA** | Dropbox | 64 | 500 | Brain Vision system | 20[*] (2) |
| **Medellin, COL** | N.A. | 58 | 1000 | NEUROSCAN SynAmps 2 | 46 |
| **Oslo, NOR** | OpenNeuro | 64 | 1024 | BioSemi ActiveTwo system | 36 |
| **Stavanger 1, NOR** | DDI | 21 | 512 | SOMNO HD eco - SOMNOmedics | 44 |
| **Stavanger 2, NOR** | EDLB | 21 | 512 | SOMNO HD eco - SOMNOmedics | 89 |

[a] Individuals with a minimum number of 20 clean 5-second-length epochs (total signal time > = 100 seconds per subject) were included in this analysis.

[*] The number of excluded subjects with shorter signals is presented in brackets.

**Supplementary Table 4. Demographic characteristics of the samples from Independent Datasets**

| Site (n) | Group (n) | Sex (n) | Median Age (years) | Age p25 (years) | Age p25 (years) |
|---|---|---|---|---|---|
| **California (n = 31)** | **HC (n = 16)** | **F (n = 9)** | 59.00 | 54.00 | 61.00 |
| | | **M (n = 7)** | 69.00 | 64.00 | 73.50 |
| | **PD (n = 15)** | **F (n = 8)** | 66.50 | 60.25 | 69.50 |
| | | **M (n = 7)** | 62.00 | 58.00 | 68.50 |
| **Chieti (n = 104)** | **AD (n = 38)** | **F (n = 23)** | 76.00 | 72.00 | 80.00 |
| | | **M (n = 15)** | 73.00 | 71.00 | 76.00 |
| | **LBD (n = 63)** | **F (n = 27)** | 76.00 | 72.00 | 80.00 |
| | | **M (n = 36)** | 77.00 | 71.75 | 80.00 |
| | **MCI_AD (n = 1)** | **F (n = 1)** | 84.00 | 84.00 | 84.00 |
| | **PD (n = 2)** | **F (n = 1)** | 68.00 | 68.00 | 68.00 |
| | | **M (n = 1)** | 71.00 | 71.00 | 71.00 |
| **Genoa (n = 77)** | **AD (n = 77)** | **F (n = 54)** | 77.00 | 74.00 | 83.00 |
| | | **M (n = 23)** | 76.00 | 74.00 | 79.50 |
| **Iowa (n = 28)** | **HC (n = 14)** | **F (n = 8)** | 69.00 | 64.75 | 72.50 |
| | | **M (n = 6)** | 72.00 | 70.00 | 78.50 |
| | **MCI_LBD (n = 2)** | **M (n = 2)** | 80.00 | 80.00 | 80.00 |
| | **PD (n = 12)** | **F (n = 8)** | 69.00 | 64.75 | 72.50 |
| | | **M (n = 4)** | 70.00 | 67.50 | 71.00 |
| **Medellin (n = 72)** | **HC (n = 36)** | **F (n = 12)** | 62.00 | 58.75 | 65.00 |
| | | **M (n = 24)** | 65.00 | 60.50 | 69.00 |
| | **MCI_LBD (n = 14)** | **F (n = 4)** | 62.00 | 60.75 | 62.25 |
| | | **M (n = 10)** | 69.00 | 65.75 | 71.00 |
| | **PD (n = 22)** | **F (n = 8)** | 62.50 | 54.75 | 66.75 |
| | | **M (n = 14)** | 63.00 | 56.50 | 65.75 |
| **Newcastle (n = 57)** | **AD (n = 32)** | **F (n = 10)** | 79.50 | 74.50 | 85.75 |
| | | **M (n = 22)** | 78.00 | 71.25 | 80.00 |
| | **LBD (n = 25)** | **F (n = 5)** | 81.00 | 81.00 | 81.00 |
| | | **M (n = 20)** | 75.50 | 70.75 | 78.00 |
| **Oslo (n = 35)** | **HC (n = 35)** | **F (n = 22)** | 52.00 | 50.00 | 60.00 |
| | | **M (n = 13)** | 54.00 | 46.00 | 65.00 |
| **Paris (n = 74)** | **MCI_AD (n = 58)** | **F (n = 34)** | 73.01 | 66.38 | 78.26 |
| | | **M (n = 24)** | 67.66 | 60.23 | 77.11 |
| | **MCI_LBD (n = 16)** | **F (n = 3)** | 70.00 | 68.50 | 73.00 |
| | | **M (n = 13)** | 72.00 | 62.00 | 75.00 |
| **Stavanger (n = 36)** | **AD (n = 3)** | **F (n = 3)** | 74.20 | 70.90 | 79.05 |
| | **HC (n = 5)** | **F (n = 2)** | 76.00 | 70.50 | 81.50 |
| | | **M (n = 3)** | 80.00 | 63.00 | 83.00 |
| | **LBD (n = 7)** | **F (n = 2)** | 77.75 | 77.42 | 78.08 |
| | | **M (n = 5)** | 72.50 | 71.00 | 77.00 |
| | **MCI_AD (n = 18)** | **F (n = 8)** | 75.00 | 69.75 | 78.25 |
| | | **M (n = 10)** | 75.00 | 70.50 | 77.00 |
| | **MCI_LBD (n = 2)** | **F (n = 1)** | 79.00 | 79.00 | 79.00 |
| | | **M (n = 1)** | 66.20 | 66.20 | 66.20 |
| | **PD (n = 1)** | **F (n = 1)** | 78.30 | 78.30 | 78.30 |
| **Thessaloniki (n = 87)** | **AD (n = 36)** | **F (n = 24)** | 67.00 | 60.50 | 72.25 |

| | | | | | |
|---|---|---|---|---|---|
| | | **M (n = 12)** | 69.50 | 61.75 | 71.00 |
| | **FTD (n = 23)** | **F (n = 9)** | 67.00 | 60.00 | 71.00 |
| | | **M (n = 14)** | 63.50 | 61.25 | 69.00 |
| | **HC (n = 28)** | **F (n = 11)** | 70.00 | 64.00 | 72.00 |
| | | **M (n = 17)** | 66.00 | 63.00 | 70.00 |
| **Turku (n = 38)** | **HC (n = 19)** | **F (n = 12)** | 67.00 | 64.00 | 69.50 |
| | | **M (n = 7)** | 70.00 | 64.50 | 75.00 |
| | **PD (n = 19)** | **F (n = 11)** | 69.00 | 64.50 | 75.50 |
| | | **M (n = 8)** | 71.00 | 66.00 | 76.50 |

**HC:** Healthy Controls; **FTD:** Frontotemporal Dementia; **AD:** Alzheimer's Disease; **PD:** Parkinson's Disease; **LBD:** Lewy Body Dementia (comprising dementia in PD and Dementia with Lewy Bodies – DLB); **MCI-LBD:** Mild Cognitive Impairment in Lewy Body Dementia (comprising MCI in PD and MCI with reported Lewy Bodies pathology); **MCI-AD:** Mild Cognitive Impairment with reported AD pathology (or without Lewy Bodies); **F:** Female; **M:** Male; **p25 & p 75:** percentile 25 & percentile 75 of Age.

**Supplementary Table 5. Age-related differences in the Pooled sample across sites**

| Group A | Group B | mean(A) | mean(B) | Difference | S.E. | T-value | d.f. | p-value | Hedge's G |
|---|---|---|---|---|---|---|---|---|---|
| **California** | **Chieti** | 63.387 | 75.212 | -11.824 | 1.723 | -6.864 | 41.228 | **< 0.001** | -1.605 |
| | **Genoa** | 63.387 | 77.247 | -13.860 | 1.741 | -7.963 | 42.682 | **< 0.001** | -1.941 |
| | **Iowa** | 63.387 | 70.500 | -7.113 | 2.258 | -3.150 | 56.763 | 0.084 | -0.809 |
| | **Medellin** | 63.387 | 63.403 | -0.016 | 1.795 | -0.009 | 47.481 | 1.000 | -0.002 |
| | **Newcastle** | 63.387 | 76.421 | -13.034 | 1.842 | -7.076 | 51.098 | **< 0.001** | -1.673 |
| | **Oslo** | 63.387 | 54.800 | 8.587 | 2.051 | 4.188 | 59.906 | **0.004** | 1.029 |
| | **Paris** | 63.387 | 70.703 | -7.316 | 1.967 | -3.720 | 63.251 | **0.017** | -0.751 |
| | **Stavanger** | 63.387 | 74.306 | -10.918 | 2.040 | -5.353 | 59.993 | **< 0.001** | -1.310 |
| | **Thessaloniki** | 63.387 | 66.103 | -2.716 | 1.774 | -1.532 | 45.767 | 0.901 | -0.347 |
| | **Turku** | 63.387 | 68.553 | -5.166 | 1.959 | -2.636 | 56.980 | 0.255 | -0.645 |
| **Chieti** | **Genoa** | 75.212 | 77.247 | -2.035 | 0.979 | -2.080 | 170.851 | 0.593 | -0.307 |
| | **Iowa** | 75.212 | 70.500 | 4.712 | 1.740 | 2.707 | 36.889 | 0.235 | 0.650 |
| | **Medellin** | 75.212 | 63.403 | 11.809 | 1.072 | 11.015 | 148.751 | **< 0.001** | 1.694 |
| | **Newcastle** | 75.212 | 76.421 | -1.210 | 1.149 | -1.053 | 112.139 | 0.993 | -0.174 |
| | **Oslo** | 75.212 | 54.800 | 20.412 | 1.460 | 13.976 | 53.221 | **< 0.001** | 2.881 |
| | **Paris** | 75.212 | 70.703 | 4.508 | 1.340 | 3.364 | 120.138 | **0.039** | 0.542 |
| | **Stavanger** | 75.212 | 74.306 | 0.906 | 1.445 | 0.627 | 55.298 | 1.000 | 0.128 |
| | **Thessaloniki** | 75.212 | 66.103 | 9.108 | 1.036 | 8.791 | 177.297 | **< 0.001** | 1.281 |
| | **Turku** | 75.212 | 68.553 | 6.659 | 1.329 | 5.009 | 63.727 | **< 0.001** | 0.961 |
| **Genoa** | **Iowa** | 77.247 | 70.500 | 6.747 | 1.758 | 3.838 | 38.195 | **0.017** | 0.968 |
| | **Medellin** | 77.247 | 63.403 | 13.844 | 1.101 | 12.577 | 141.753 | **< 0.001** | 2.060 |
| | **Newcastle** | 77.247 | 76.421 | 0.826 | 1.176 | 0.702 | 112.308 | 1.000 | 0.124 |
| | **Oslo** | 77.247 | 54.800 | 22.447 | 1.482 | 15.150 | 55.419 | **< 0.001** | 3.310 |
| | **Paris** | 77.247 | 70.703 | 6.544 | 1.363 | 4.800 | 121.914 | **< 0.001** | 0.784 |
| | **Stavanger** | 77.247 | 74.306 | 2.941 | 1.467 | 2.005 | 57.569 | 0.646 | 0.433 |
| | **Thessaloniki** | 77.247 | 66.103 | 11.143 | 1.066 | 10.456 | 161.737 | **< 0.001** | 1.612 |
| | **Turku** | 77.247 | 68.553 | 8.694 | 1.353 | 6.428 | 66.302 | **< 0.001** | 1.320 |
| **Iowa** | **Medellin** | 70.500 | 63.403 | 7.097 | 1.812 | 3.917 | 42.486 | **0.013** | 0.937 |
| | **Newcastle** | 70.500 | 76.421 | -5.921 | 1.858 | -3.186 | 45.824 | 0.082 | -0.777 |
| | **Oslo** | 70.500 | 54.800 | 15.700 | 2.065 | 7.601 | 55.115 | **< 0.001** | 1.925 |
| | **Paris** | 70.500 | 70.703 | -0.203 | 1.982 | -0.103 | 56.843 | 1.000 | -0.021 |
| | **Stavanger** | 70.500 | 74.306 | -3.806 | 2.055 | -1.852 | 55.078 | 0.744 | -0.467 |
| | **Thessaloniki** | 70.500 | 66.103 | 4.397 | 1.791 | 2.455 | 40.926 | 0.359 | 0.570 |
| | **Turku** | 70.500 | 68.553 | 1.947 | 1.975 | 0.986 | 51.791 | 0.995 | 0.250 |
| **Medellin** | **Newcastle** | 63.403 | 76.421 | -13.018 | 1.255 | -10.375 | 120.707 | **< 0.001** | -1.827 |
| | **Oslo** | 63.403 | 54.800 | 8.603 | 1.545 | 5.568 | 62.979 | **< 0.001** | 1.170 |
| | **Paris** | 63.403 | 70.703 | -7.300 | 1.432 | -5.099 | 132.000 | **< 0.001** | -0.836 |
| | **Stavanger** | 63.403 | 74.306 | -10.903 | 1.531 | -7.123 | 65.431 | **< 0.001** | -1.482 |
| | **Thessaloniki** | 63.403 | 66.103 | -2.701 | 1.152 | -2.344 | 153.387 | 0.410 | -0.370 |
| | **Turku** | 63.403 | 68.553 | -5.150 | 1.422 | -3.622 | 75.664 | **0.021** | -0.720 |
| **Newcastle** | **Oslo** | 76.421 | 54.800 | 21.621 | 1.599 | 13.518 | 67.364 | **< 0.001** | 2.938 |
| | **Paris** | 76.421 | 70.703 | 5.718 | 1.490 | 3.837 | 128.117 | **0.009** | 0.643 |
| | **Stavanger** | 76.421 | 74.306 | 2.115 | 1.586 | 1.334 | 69.787 | 0.959 | 0.287 |
| | **Thessaloniki** | 76.421 | 66.103 | 10.318 | 1.224 | 8.428 | 123.688 | **< 0.001** | 1.415 |
| | **Turku** | 76.421 | 68.553 | 7.868 | 1.481 | 5.314 | 79.229 | **< 0.001** | 1.105 |

| | | | | | | | | | |
|---|---|---|---|---|---|---|---|---|---|
| **Oslo** | **Paris** | 54.800 | 70.703 | -15.903 | 1.742 | -9.131 | 84.862 | **< 0.001** | -1.695 |
| | **Stavanger** | 54.800 | 74.306 | -19.506 | 1.824 | -10.694 | 68.947 | **< 0.001** | -2.511 |
| | **Thessaloniki** | 54.800 | 66.103 | -11.303 | 1.520 | -7.435 | 60.631 | **< 0.001** | -1.505 |
| | **Turku** | 54.800 | 68.553 | -13.753 | 1.733 | -7.933 | 69.138 | **< 0.001** | -1.845 |
| **Paris** | **Stavanger** | 70.703 | 74.306 | -3.602 | 1.729 | -2.083 | 87.735 | 0.592 | -0.384 |
| | **Thessaloniki** | 70.703 | 66.103 | 4.600 | 1.405 | 3.274 | 132.298 | 0.050 | 0.528 |
| | **Turku** | 70.703 | 68.553 | 2.151 | 1.633 | 1.317 | 98.901 | 0.964 | 0.234 |
| **Stavanger** | **Thessaloniki** | 74.306 | 66.103 | 8.202 | 1.506 | 5.447 | 63.047 | **< 0.001** | 1.091 |
| | **Turku** | 74.306 | 68.553 | 5.753 | 1.721 | 3.343 | 70.684 | **0.048** | 0.771 |
| **Thessaloniki** | **Turku** | 66.103 | 68.553 | -2.449 | 1.395 | -1.756 | 73.318 | 0.801 | -0.334 |

Games-Howell test estimates are presented. Statistically significant p-values are shown in bold.

**S.E:** Standard Error**; d.f:** Adjusted degrees of freedom.

**Supplementary Table 6. Age-related differences in the Pooled sample by Diagnosis**

| Group A | Group B | mean(A) | mean(B) | Difference | S.E. | T-value | d.f. | p-value | Hedge's G |
|---|---|---|---|---|---|---|---|---|---|
| **AD** | **FTD** | 74.428 | 63.652 | 10.776 | 1.818 | 5.929 | 27.748 | **< 0.001** | 1.304 |
| | **HC** | 74.428 | 63.693 | 10.736 | 0.963 | 11.152 | 307.066 | **< 0.001** | 1.229 |
| | **LBD** | 74.428 | 75.829 | -1.401 | 0.859 | -1.630 | 247.302 | 0.663 | -0.185 |
| | **MCI_AD** | 74.428 | 71.754 | 2.675 | 1.253 | 2.135 | 124.306 | 0.339 | 0.308 |
| | **MCI_LBD** | 74.428 | 69.241 | 5.187 | 1.533 | 3.383 | 45.954 | **0.023** | 0.628 |
| | **PD** | 74.428 | 65.849 | 8.579 | 1.184 | 7.246 | 122.175 | **< 0.001** | 1.027 |
| **FTD** | **HC** | 63.652 | 63.693 | -0.041 | 1.871 | -0.022 | 31.052 | 1.000 | -0.004 |
| | **LBD** | 63.652 | 75.829 | -12.177 | 1.820 | -6.690 | 27.846 | **< 0.001** | -1.876 |
| | **MCI_AD** | 63.652 | 71.754 | -8.102 | 2.036 | -3.980 | 41.703 | **0.005** | -0.861 |
| | **MCI_LBD** | 63.652 | 69.241 | -5.589 | 2.219 | -2.518 | 47.364 | 0.176 | -0.671 |
| | **PD** | 63.652 | 65.849 | -2.197 | 1.994 | -1.102 | 38.750 | 0.924 | -0.256 |
| **HC** | **LBD** | 63.693 | 75.829 | -12.137 | 0.967 | -12.546 | 245.686 | **< 0.001** | -1.482 |
| | **MCI_AD** | 63.693 | 71.754 | -8.061 | 1.329 | -6.065 | 147.401 | **< 0.001** | -0.855 |
| | **MCI_LBD** | 63.693 | 69.241 | -5.548 | 1.596 | -3.476 | 53.383 | **0.017** | -0.608 |
| | **PD** | 63.693 | 65.849 | -2.156 | 1.265 | -1.705 | 146.586 | 0.614 | -0.237 |
| **LBD** | **MCI_AD** | 75.829 | 71.754 | 4.076 | 1.256 | 3.244 | 121.057 | **0.025** | 0.519 |
| | **MCI_LBD** | 75.829 | 69.241 | 6.588 | 1.536 | 4.288 | 46.016 | **0.002** | 0.989 |
| | **PD** | 75.829 | 65.849 | 9.980 | 1.188 | 8.402 | 118.124 | **< 0.001** | 1.381 |
| **MCI_AD** | **MCI_LBD** | 71.754 | 69.241 | 2.513 | 1.786 | 1.406 | 73.435 | 0.797 | 0.270 |
| | **PD** | 71.754 | 65.849 | 5.904 | 1.497 | 3.943 | 145.835 | **0.002** | 0.642 |
| **MCI_LBD** | **PD** | 69.241 | 65.849 | 3.392 | 1.739 | 1.951 | 67.748 | 0.455 | 0.398 |

Games-Howell test estimates are presented. Statistically significant differences are shown in bold.

**HC:** Healthy Controls; **FTD:** Frontotemporal Dementia; **AD:** Alzheimer's Disease; **PD:** Parkinson's Disease; **LBD:** Lewy Body Dementia (comprising dementia in PD and Dementia with Lewy Bodies – DLB); **MCI-LBD:** Mild Cognitive Impairment in Lewy Body Dementia (comprising MCI in PD and MCI with reported Lewy Bodies pathology); **MCI-AD:** Mild Cognitive Impairment with reported AD pathology (or without Lewy Bodies); **S.E:** Standard Error**; d.f:** Adjusted degrees of freedom.

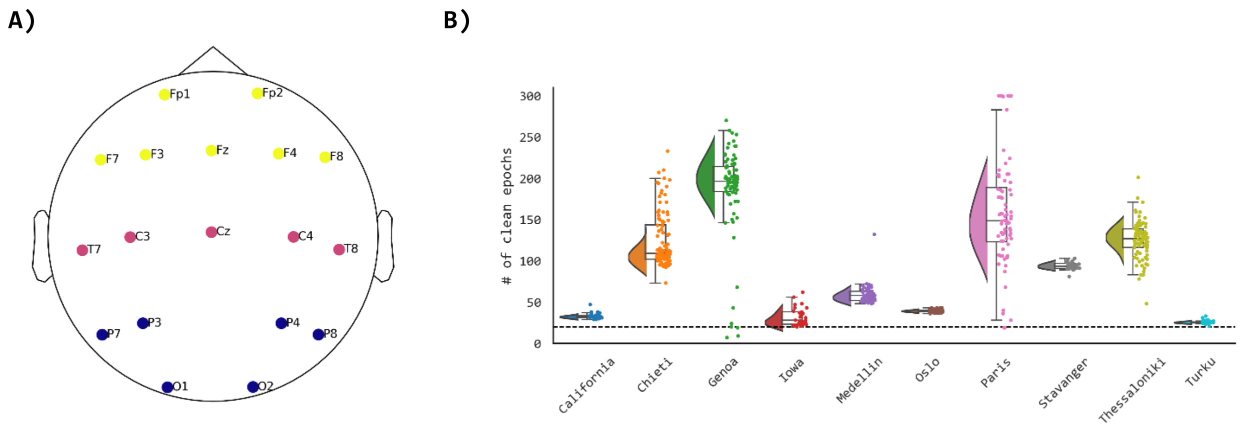


**Supplementary Figure 1. Common electrode positions across sites and signal length after preprocessing.** (A) Topoplot depicts the common electrode positions across sites following the international 10-20 system disposition. Colors illustrate the frontal (yellow), central (pink), and posterior (blue) regions of interest (ROI). Further analyses were performed in the posterior ROI. (B) Raincloud plots show the number of clean epochs (5 seconds-length) across sites. Dots represent the number of non-artifactual epochs in each recording. The horizontal dashed line marks the signals included in the study (i.e., those with 20 or more epochs, 100 seconds, after preprocessing).

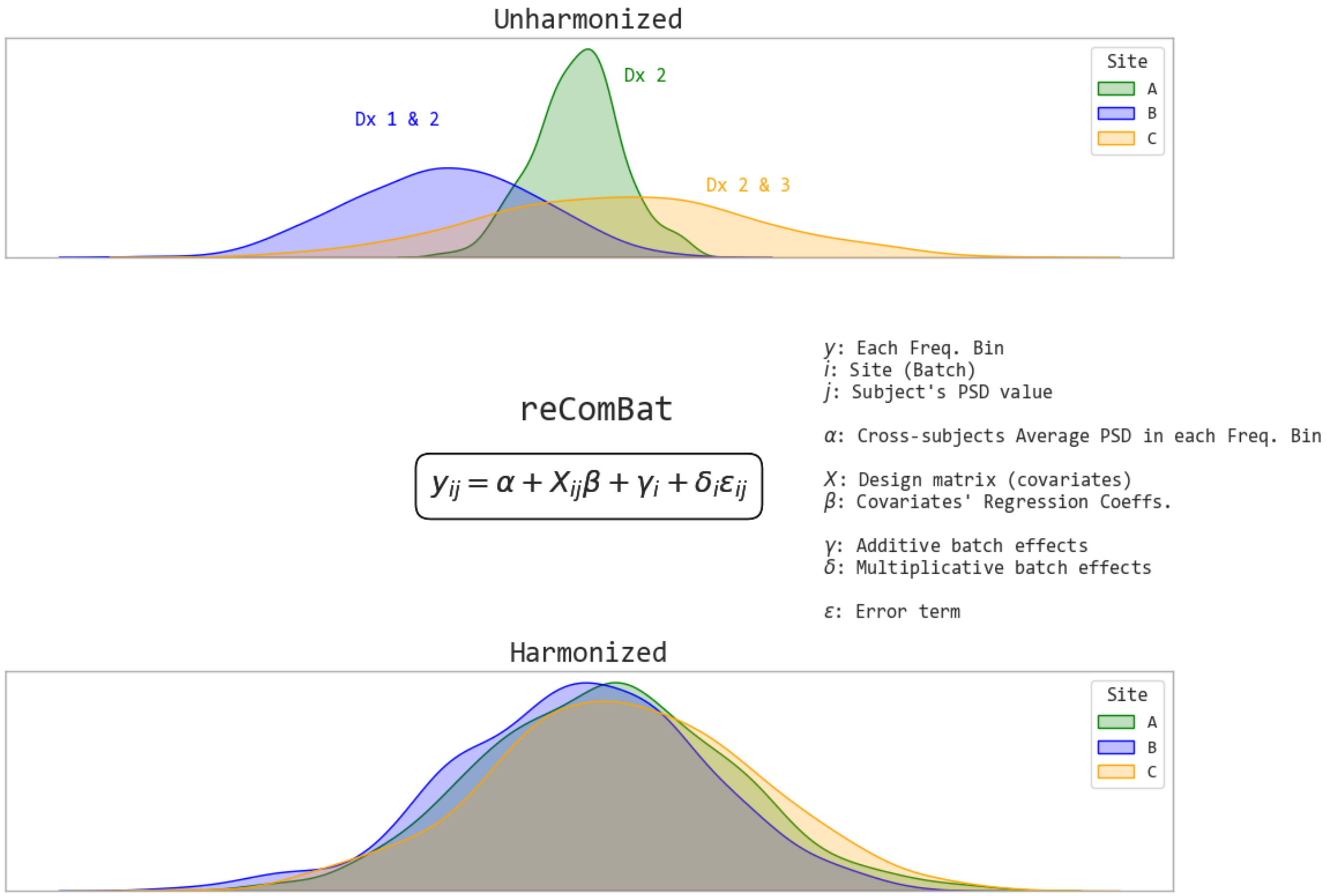


**Supplementary Figure 2. Graphical representation of the reComBat Batch Harmonization model.** Histograms on top illustrate simulated data distributions from a single feature (each frequency bin, in this study) across three sites (batches). The subjects on each batch were sampled from populations with three possible diagnoses (Dx). Of note, site A (green) only included subjects with a single diagnosis (Dx2), resulting in a singular design matrix. The reComBat harmonization model assumes that additive and multiplicative batch effects can be estimated and regressed out of the feature values while preserving the variance explained by biological covariates of interest. Thus, reComBat returns the re-scaled and re-centered features with statistically comparable distributions (bottom). If the design matrix is singular, reComBat uses regularization methods to solve it. **PSD:** Power spectrum. **Freq:** Frequency. **Coeffs:** Coefficients.

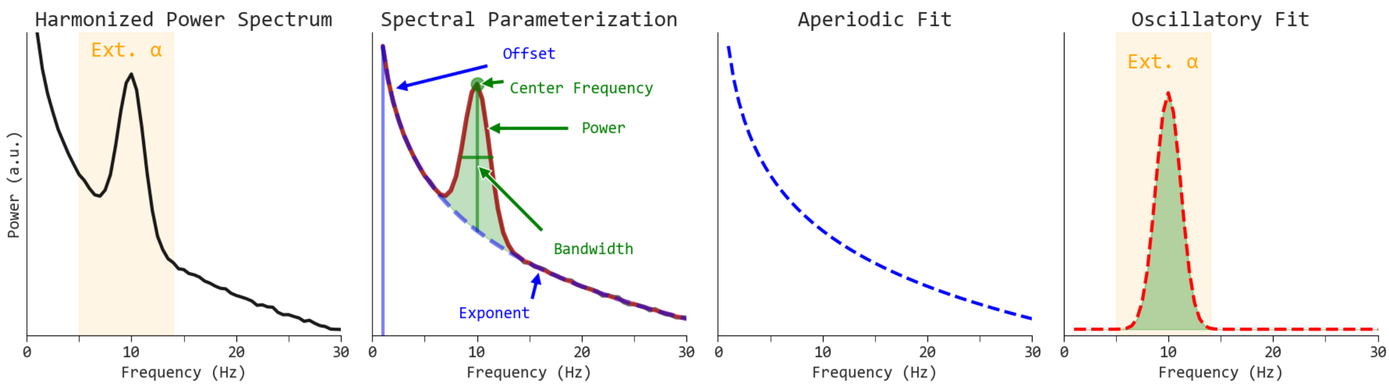


**Supplementary Figure 3. Graphical representation of the spectral parameterization of the power spectrum.** From left to right, the batch harmonized power spectrum is taken as input for spectral parameterization with the Fitting Oscillations and One-Over Frequency (FOOOF) algorithm. FOOOF decomposes the power spectrum (black) into oscillatory (red) and aperiodic activity (blue) vectors. From these vectors, FOOOF computes aperiodic parameters (exponent, offset). In addition, oscillatory parameters (center frequency, power, bandwidth) are estimated within a particular frequency band (in this study, the Extended alpha band, yellow).

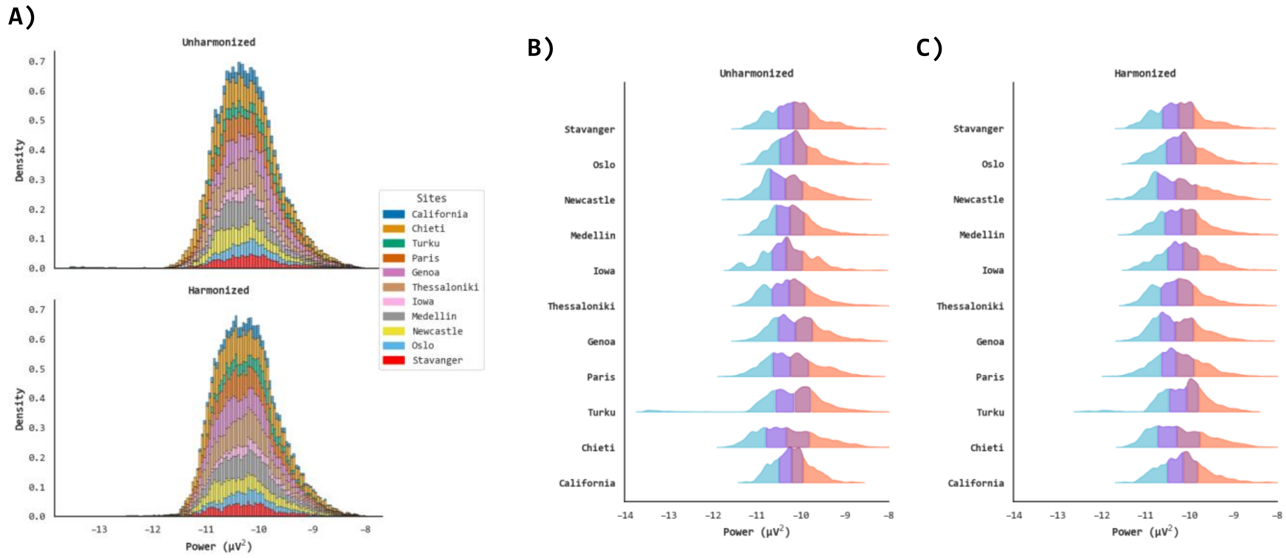


**Supplementary Figure 4. Qualitative visualization of batch effects harmonization - Univariate power spectrum plots.** Stacked histograms of power spectrum by site before and after harmonization (A); each color represents a site. Ridgeline plots (Joyplots) contrast the power spectrum distributions across sites in the unharmonized (B) and harmonized (C) datasets; blue-purple-pink-red colors represent quantiles.

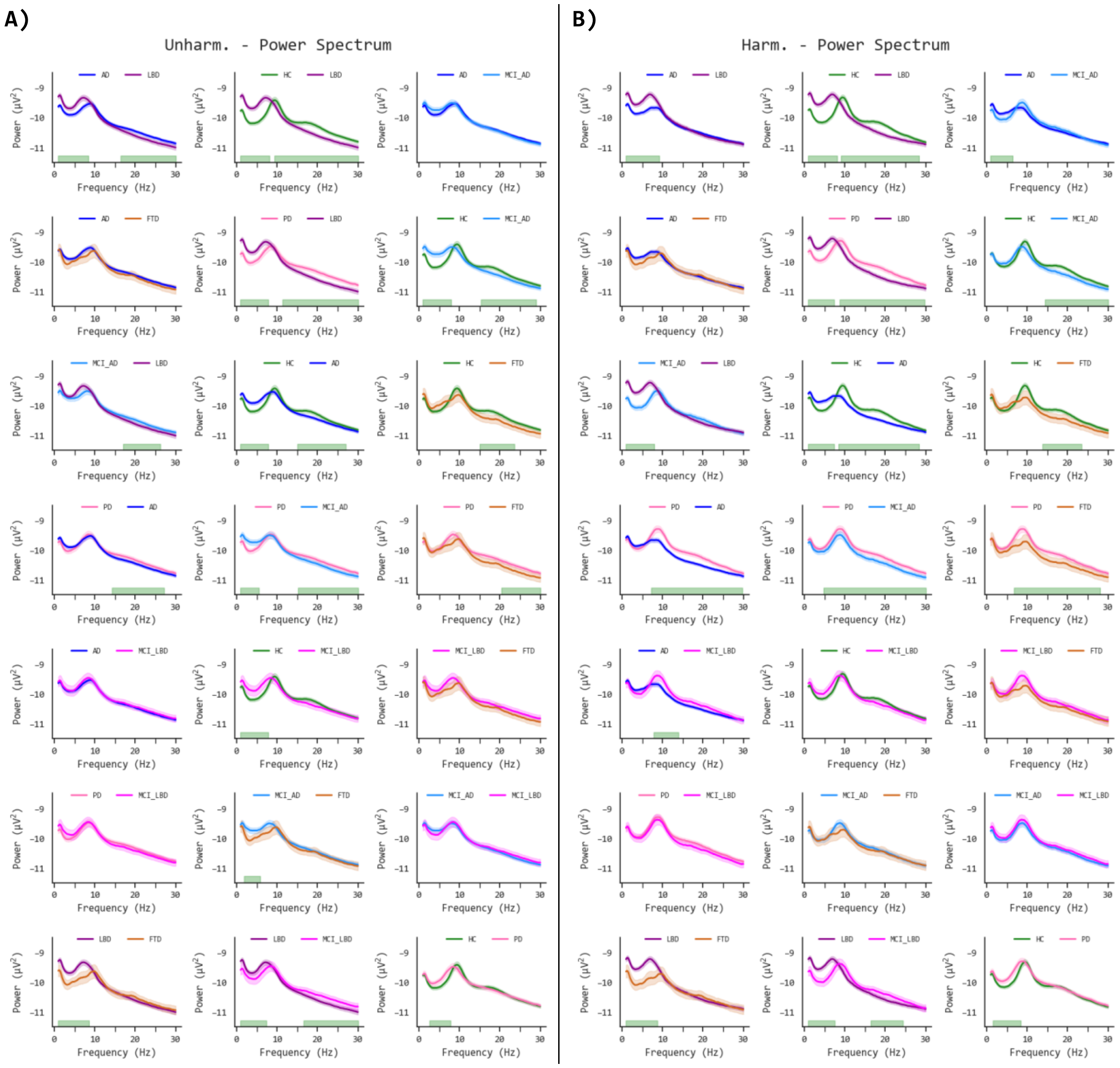


**Supplementary Figure 5. Pairwise comparisons of the unharmonized and batch harmonized power spectrum across neurodegenerative diseases (NDDs). (A)** Unharmonized mean power spectrum (lines) and 95 % standard error (shaded area); green regions on the x-axis bottom represent significant p values ($p < 0.05$) on mass univariate permutation F-value tests (1000 permutations) clustered on frequencies. **(B)** Harmonized mean power spectrum (lines) and 95 % standard error (shaded area); green regions on the x-axis bottom represent significant clusters ($p < 0.05$). **HC:** Healthy Controls; **FTD:** Frontotemporal Dementia; **AD:** Alzheimer's Disease; **PD:** Parkinson's Disease; **LBD:** Lewy Body Dementia (comprising dementia in PD and Dementia with Lewy Bodies – DLB); **MCI-LBD:** Mild Cognitive Impairment in Lewy Body Dementia (comprising MCI in PD and MCI with reported Lewy Bodies pathology); **MCI-AD:** Mild Cognitive Impairment with reported AD pathology (or without Lewy Bodies).

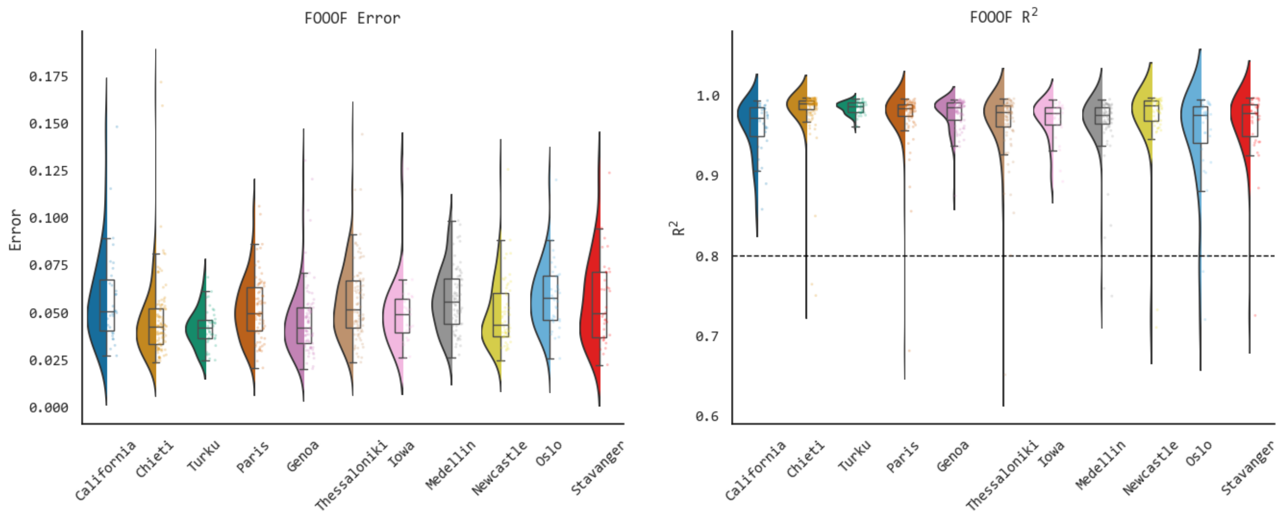


**Supplementary Figure 6. Performance of the Spectral parameterization fitting.** Raincloud plots show the mean absolute error and R-squared values obtained from the Fitting Oscillations and One-Over-Frequency (FOOOF) model. The horizontal dashed line indicates the minimum cutoff for the inclusion of FOOOF derivatives in subsequent analyses.

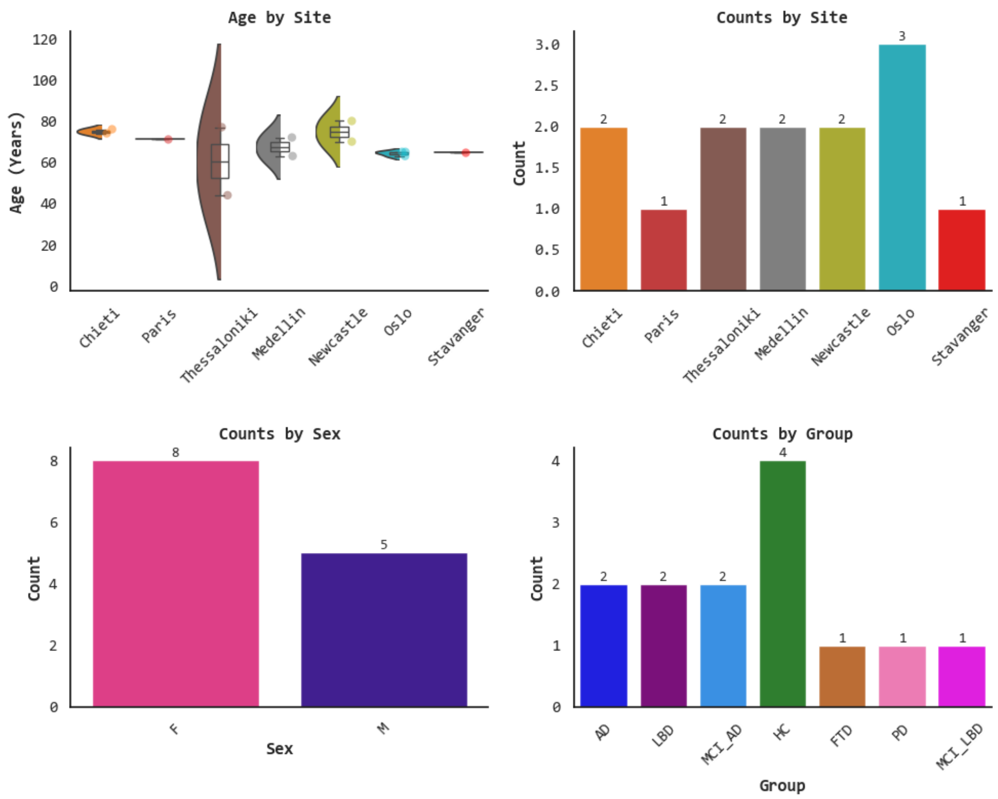


**Supplementary Figure 7. Demographic characteristics of subjects with suboptimal spectral parameterization fitting (n = 13).** Raincloud plots show age by site. Barplots show absolute frequency by site, sex, and group. **HC:** Healthy Controls; **FTD:** Frontotemporal Dementia; **AD:** Alzheimer's Disease; **PD:** Parkinson's Disease; **LBD:** Lewy Body Dementia (comprising dementia in PD and Dementia with Lewy Bodies – DLB); **MCI-LBD:** Mild Cognitive Impairment in Lewy Body Dementia (comprising MCI in PD and MCI with reported Lewy Bodies pathology); **MCI-AD:** Mild Cognitive Impairment with reported AD pathology (or without Lewy Bodies); **F:** Female; **M:** Male.

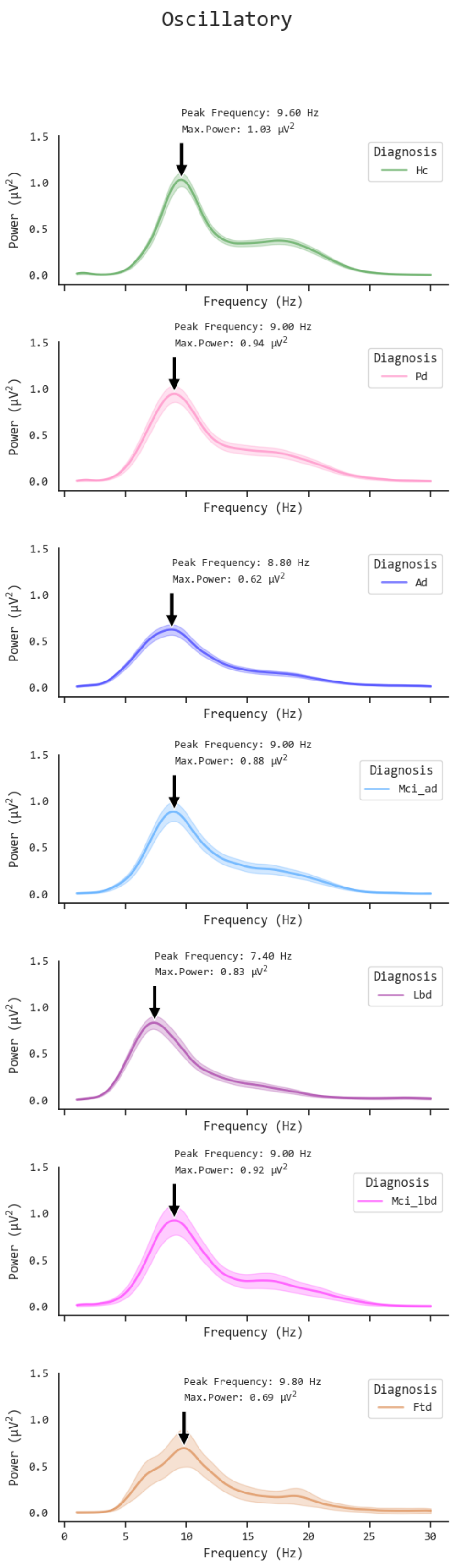


**Supplementary Figure 8. Oscillatory fitting vectors across neurodegenerative diseases (NDDs).** Each subplot represents the mean oscillatory fit (lines) and 95 % standard error (shaded area) across NDDs. The peak frequency and maximum peak power values are annotated. **HC:** Healthy Controls; **FTD:** Frontotemporal Dementia; **AD:** Alzheimer's Disease; **PD:** Parkinson's Disease; **LBD:** Lewy Body Dementia (comprising dementia in PD and Dementia with Lewy Bodies – DLB); **MCI-LBD:** Mild Cognitive Impairment in Lewy Body Dementia (comprising MCI in PD and MCI with reported Lewy Bodies pathology); **MCI-AD:** Mild Cognitive Impairment with reported AD pathology (or without Lewy Bodies).

Ap. Exponent

| Variable | | Mean A | SD A | Mean B | SD B |
|---|---|---|---|---|---|
| LBD VS FTD | 0.43( 0.22 to 0.64) | 1.302 | 0.391 | 0.87 | 0.486 |
| AD VS PD | 0.19( 0.11 to 0.28) | 0.962 | 0.335 | 0.767 | 0.322 |
| AD VS MCI_LBD | 0.11(-0.06 to 0.27) | 0.962 | 0.335 | 0.849 | 0.478 |
| AD VS FTD | 0.09(-0.11 to 0.30) | 0.962 | 0.335 | 0.87 | 0.486 |
| MCI_AD VS PD | 0.04(-0.09 to 0.17) | 0.804 | 0.445 | 0.767 | 0.322 |
| MCI_LBD VS FTD | -0.02(-0.28 to 0.25) | 0.849 | 0.478 | 0.87 | 0.486 |
| MCI_AD VS MCI_LBD | -0.04(-0.25 to 0.15) | 0.804 | 0.445 | 0.849 | 0.478 |
| MCI_AD VS FTD | -0.07(-0.30 to 0.14) | 0.804 | 0.445 | 0.87 | 0.486 |
| PD VS MCI_LBD | -0.08(-0.30 to 0.08) | 0.767 | 0.322 | 0.849 | 0.478 |
| HC VS PD | -0.10(-0.18 to -0.01) | 0.671 | 0.276 | 0.767 | 0.322 |
| PD VS FTD | -0.10(-0.35 to 0.09) | 0.767 | 0.322 | 0.87 | 0.486 |
| HC VS MCI_AD | -0.13(-0.23 to -0.03) | 0.671 | 0.276 | 0.804 | 0.445 |
| MCI_AD VS AD | -0.16(-0.27 to -0.05) | 0.804 | 0.445 | 0.962 | 0.335 |
| HC VS MCI_LBD | -0.18(-0.37 to -0.04) | 0.671 | 0.276 | 0.849 | 0.478 |
| HC VS FTD | -0.20(-0.41 to -0.00) | 0.671 | 0.276 | 0.87 | 0.486 |
| HC VS AD | -0.29(-0.36 to -0.22) | 0.671 | 0.276 | 0.962 | 0.335 |
| AD VS LBD | -0.34(-0.43 to -0.25) | 0.962 | 0.335 | 1.302 | 0.391 |
| MCI_LBD VS LBD | -0.45(-0.63 to -0.27) | 0.849 | 0.478 | 1.302 | 0.391 |
| MCI_AD VS LBD | -0.50(-0.64 to -0.38) | 0.804 | 0.445 | 1.302 | 0.391 |
| PD VS LBD | -0.54(-0.64 to -0.43) | 0.767 | 0.322 | 1.302 | 0.391 |
| HC VS LBD | -0.63(-0.73 to -0.54) | 0.671 | 0.276 | 1.302 | 0.391 |

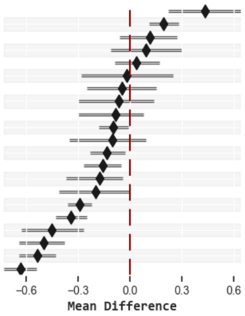


Ap. Offset

| Variable | | Mean A | SD A | Mean B | SD B |
|---|---|---|---|---|---|
| LBD VS FTD | 0.60( 0.33 to 0.85) | -8.973 | 0.44 | -9.57 | 0.6 |
| AD VS FTD | 0.15(-0.09 to 0.40) | -9.422 | 0.42 | -9.57 | 0.6 |
| AD VS PD | 0.11(-0.01 to 0.21) | -9.422 | 0.42 | -9.529 | 0.395 |
| AD VS MCI_LBD | 0.09(-0.12 to 0.27) | -9.422 | 0.42 | -9.516 | 0.551 |
| MCI_LBD VS FTD | 0.05(-0.26 to 0.36) | -9.516 | 0.551 | -9.57 | 0.6 |
| PD VS FTD | 0.04(-0.25 to 0.27) | -9.529 | 0.395 | -9.57 | 0.6 |
| PD VS MCI_LBD | -0.01(-0.24 to 0.18) | -9.529 | 0.395 | -9.516 | 0.551 |
| MCI_AD VS FTD | -0.06(-0.33 to 0.20) | -9.629 | 0.475 | -9.57 | 0.6 |
| HC VS MCI_AD | -0.08(-0.20 to 0.06) | -9.707 | 0.37 | -9.629 | 0.475 |
| MCI_AD VS PD | -0.10(-0.24 to 0.06) | -9.629 | 0.475 | -9.529 | 0.395 |
| MCI_AD VS MCI_LBD | -0.11(-0.35 to 0.10) | -9.629 | 0.475 | -9.516 | 0.551 |
| HC VS FTD | -0.14(-0.39 to 0.11) | -9.707 | 0.37 | -9.57 | 0.6 |
| HC VS PD | -0.18(-0.29 to -0.06) | -9.707 | 0.37 | -9.529 | 0.395 |
| HC VS MCI_LBD | -0.19(-0.41 to -0.01) | -9.707 | 0.37 | -9.516 | 0.551 |
| MCI_AD VS AD | -0.21(-0.32 to -0.08) | -9.629 | 0.475 | -9.422 | 0.42 |
| HC VS AD | -0.28(-0.37 to -0.20) | -9.707 | 0.37 | -9.422 | 0.42 |
| AD VS LBD | -0.45(-0.56 to -0.35) | -9.422 | 0.42 | -8.973 | 0.44 |
| MCI_LBD VS LBD | -0.54(-0.74 to -0.34) | -9.516 | 0.551 | -8.973 | 0.44 |
| PD VS LBD | -0.56(-0.68 to -0.44) | -9.529 | 0.395 | -8.973 | 0.44 |
| MCI_AD VS LBD | -0.66(-0.80 to -0.52) | -9.629 | 0.475 | -8.973 | 0.44 |
| HC VS LBD | -0.73(-0.85 to -0.63) | -9.707 | 0.37 | -8.973 | 0.44 |

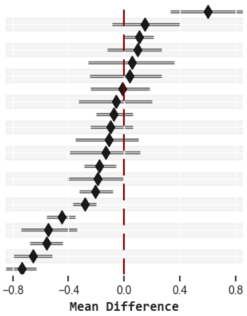


Ext. Alpha - BW

| Variable | | Mean A | SD A | Mean B | SD B |
|---|---|---|---|---|---|
| PD VS LBD | 0.25(-0.02 to 0.60) | 3.133 | 1.07 | 2.883 | 0.905 |
| PD VS FTD | 0.23(-0.40 to 0.82) | 3.133 | 1.07 | 2.904 | 1.321 |
| PD VS MCI_LBD | 0.14(-0.38 to 0.58) | 3.133 | 1.07 | 2.999 | 1.259 |
| MCI_AD VS LBD | 0.12(-0.18 to 0.41) | 3.004 | 1.076 | 2.883 | 0.905 |
| MCI_LBD VS LBD | 0.12(-0.32 to 0.65) | 2.999 | 1.259 | 2.883 | 0.905 |
| MCI_AD VS FTD | 0.10(-0.54 to 0.63) | 3.004 | 1.076 | 2.904 | 1.321 |
| MCI_LBD VS FTD | 0.10(-0.63 to 0.78) | 2.999 | 1.259 | 2.904 | 1.321 |
| AD VS LBD | 0.09(-0.18 to 0.36) | 2.969 | 1.216 | 2.883 | 0.905 |
| AD VS FTD | 0.07(-0.56 to 0.58) | 2.969 | 1.216 | 2.904 | 1.321 |
| MCI_AD VS AD | 0.03(-0.26 to 0.32) | 3.004 | 1.076 | 2.969 | 1.216 |
| MCI_AD VS MCI_LBD | 0.01(-0.53 to 0.41) | 3.004 | 1.076 | 2.999 | 1.259 |
| LBD VS FTD | -0.02(-0.63 to 0.47) | 2.883 | 0.905 | 2.904 | 1.321 |
| AD VS MCI_LBD | -0.03(-0.62 to 0.38) | 2.969 | 1.216 | 2.999 | 1.259 |
| MCI_AD VS PD | -0.13(-0.49 to 0.21) | 3.004 | 1.076 | 3.133 | 1.07 |
| HC VS LBD | -0.14(-0.39 to 0.07) | 2.737 | 0.903 | 2.883 | 0.905 |
| AD VS PD | -0.16(-0.51 to 0.14) | 2.969 | 1.216 | 3.133 | 1.07 |
| HC VS FTD | -0.17(-0.84 to 0.33) | 2.737 | 0.903 | 2.904 | 1.321 |
| HC VS AD | -0.23(-0.47 to -0.01) | 2.737 | 0.903 | 2.969 | 1.216 |
| HC VS MCI_LBD | -0.26(-0.80 to 0.15) | 2.737 | 0.903 | 2.999 | 1.259 |
| HC VS MCI_AD | -0.27(-0.57 to 0.01) | 2.737 | 0.903 | 3.004 | 1.076 |
| HC VS PD | -0.40(-0.71 to -0.13) | 2.737 | 0.903 | 3.133 | 1.07 |

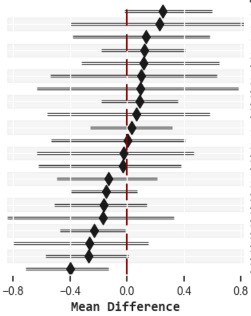


Ext. Alpha - CF

| Variable | | Mean A | SD A | Mean B | SD B |
|---|---|---|---|---|---|
| HC VS LBD | 2.23( 1.96 to 2.54) | 9.572 | 0.862 | 7.338 | 1.251 |
| PD VS LBD | 1.50( 1.12 to 1.90) | 8.843 | 1.305 | 7.338 | 1.251 |
| MCI_AD VS LBD | 1.47( 1.11 to 1.88) | 8.808 | 1.245 | 7.338 | 1.251 |
| MCI_LBD VS LBD | 1.40( 0.90 to 1.88) | 8.739 | 1.201 | 7.338 | 1.251 |
| AD VS LBD | 1.27( 0.90 to 1.59) | 8.613 | 1.708 | 7.338 | 1.251 |
| HC VS AD | 0.96( 0.69 to 1.24) | 9.572 | 0.862 | 8.613 | 1.708 |
| HC VS MCI_LBD | 0.83( 0.41 to 1.27) | 9.572 | 0.862 | 8.739 | 1.201 |
| HC VS MCI_AD | 0.76( 0.41 to 1.06) | 9.572 | 0.862 | 8.808 | 1.245 |
| HC VS PD | 0.73( 0.40 to 1.06) | 9.572 | 0.862 | 8.843 | 1.305 |
| HC VS FTD | 0.20(-0.54 to 1.02) | 9.572 | 0.862 | 9.371 | 1.902 |
| MCI_AD VS AD | 0.20(-0.14 to 0.56) | 8.808 | 1.245 | 8.613 | 1.708 |
| PD VS MCI_LBD | 0.10(-0.44 to 0.55) | 8.843 | 1.305 | 8.739 | 1.201 |
| MCI_AD VS MCI_LBD | 0.07(-0.48 to 0.55) | 8.808 | 1.245 | 8.739 | 1.201 |
| MCI_AD VS PD | -0.03(-0.45 to 0.38) | 8.808 | 1.245 | 8.843 | 1.305 |
| AD VS MCI_LBD | -0.13(-0.62 to 0.34) | 8.613 | 1.708 | 8.739 | 1.201 |
| AD VS PD | -0.23(-0.62 to 0.17) | 8.613 | 1.708 | 8.843 | 1.305 |
| PD VS FTD | -0.53(-1.38 to 0.30) | 8.843 | 1.305 | 9.371 | 1.902 |
| MCI_AD VS FTD | -0.56(-1.44 to 0.16) | 8.808 | 1.245 | 9.371 | 1.902 |
| MCI_LBD VS FTD | -0.63(-1.55 to 0.26) | 8.739 | 1.201 | 9.371 | 1.902 |
| AD VS FTD | -0.76(-1.60 to 0.05) | 8.613 | 1.708 | 9.371 | 1.902 |
| LBD VS FTD | -2.03(-2.83 to -1.20) | 7.338 | 1.251 | 9.371 | 1.902 |

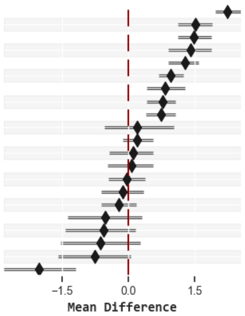


Ext. Alpha - PW

| Variable | | Mean A | SD A | Mean B | SD B |
|---|---|---|---|---|---|
| HC VS AD | 0.33( 0.23 to 0.41) | 1.158 | 0.469 | 0.826 | 0.383 |
| MCI_AD VS AD | 0.21( 0.11 to 0.33) | 1.04 | 0.433 | 0.826 | 0.383 |
| HC VS FTD | 0.20( 0.02 to 0.35) | 1.158 | 0.469 | 0.96 | 0.358 |
| HC VS LBD | 0.17( 0.07 to 0.27) | 1.158 | 0.469 | 0.983 | 0.366 |
| MCI_LBD VS FTD | 0.16(-0.06 to 0.37) | 1.118 | 0.436 | 0.96 | 0.358 |
| MCI_LBD VS LBD | 0.14(-0.02 to 0.33) | 1.118 | 0.436 | 0.983 | 0.366 |
| HC VS MCI_AD | 0.12(-0.01 to 0.23) | 1.158 | 0.469 | 1.04 | 0.433 |
| HC VS PD | 0.10(-0.02 to 0.23) | 1.158 | 0.469 | 1.059 | 0.381 |
| PD VS FTD | 0.10(-0.08 to 0.26) | 1.059 | 0.381 | 0.96 | 0.358 |
| MCI_AD VS FTD | 0.08(-0.10 to 0.25) | 1.04 | 0.433 | 0.96 | 0.358 |
| PD VS LBD | 0.08(-0.04 to 0.19) | 1.059 | 0.381 | 0.983 | 0.366 |
| MCI_AD VS LBD | 0.06(-0.06 to 0.18) | 1.04 | 0.433 | 0.983 | 0.366 |
| HC VS MCI_LBD | 0.04(-0.13 to 0.21) | 1.158 | 0.469 | 1.118 | 0.436 |
| LBD VS FTD | 0.02(-0.15 to 0.19) | 0.983 | 0.366 | 0.96 | 0.358 |
| MCI_AD VS PD | -0.02(-0.14 to 0.11) | 1.04 | 0.433 | 1.059 | 0.381 |
| PD VS MCI_LBD | -0.06(-0.22 to 0.12) | 1.059 | 0.381 | 1.118 | 0.436 |
| MCI_AD VS MCI_LBD | -0.08(-0.25 to 0.13) | 1.04 | 0.433 | 1.118 | 0.436 |
| AD VS FTD | -0.13(-0.29 to 0.02) | 0.826 | 0.383 | 0.96 | 0.358 |
| AD VS LBD | -0.16(-0.25 to -0.07) | 0.826 | 0.383 | 0.983 | 0.366 |
| AD VS PD | -0.23(-0.34 to -0.13) | 0.826 | 0.383 | 1.059 | 0.381 |
| AD VS MCI_LBD | -0.29(-0.45 to -0.13) | 0.826 | 0.383 | 1.118 | 0.436 |

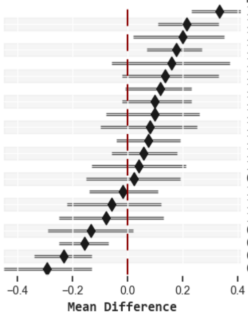


**Supplementary Figure 9. Descriptive comparisons of Spectral parameters across neurodegenerative diseases (NDDs).** Forest plots (lines) present the results of mean differences across NDDs for each spectral parameter. Mean difference was estimated and bootstrapped (1000 iterations) 95 % confidence intervals were computed. Descriptive statistics (mean and standard deviation) are presented for each pair of groups (A vs B). **HC:** Healthy Controls; **FTD:** Frontotem poral Dementia; **AD:** Alzheimer's Disease; **PD:** Parkinson's Disease; **LBD:** Lewy Body Dementia (comprising dementia in PD and Dementia with Lewy Bodies – DLB); **MCI-LBD:** Mild Cognitive Impairment in Lewy Body Dementia (comprising MCI in PD and MCI with reported Lewy Bodies pathology); **MCI-AD:** Mild Cognitive Impairment with reported AD pathology (or without Lewy Bodies).

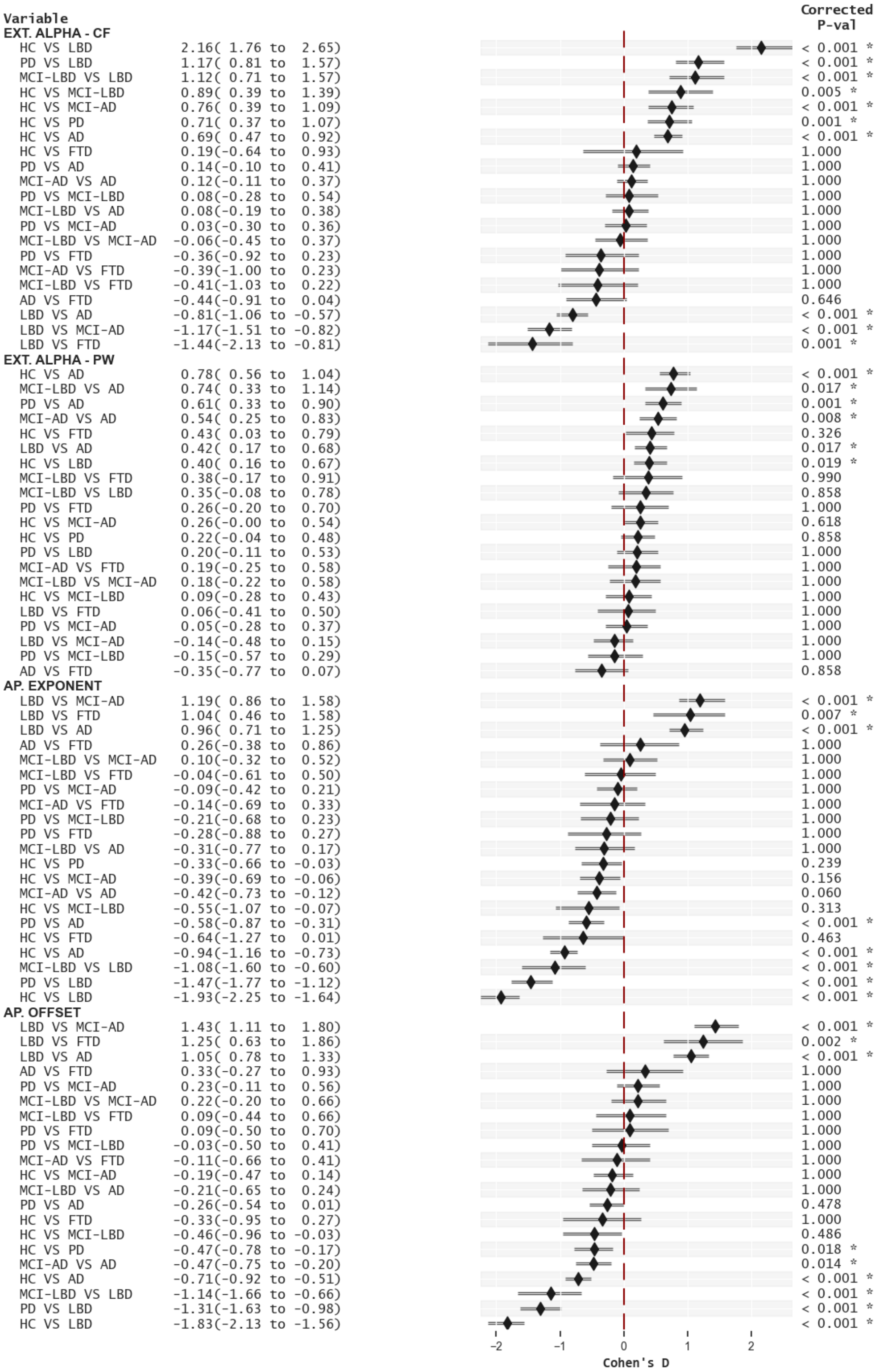


**Supplementary Figure 10. Effect Size - Comparisons of Spectral parameters across neurodegenerative diseases (NDDs).** Forest plots (lines) present the results of pairwise t-tests across NDDs. Cohen's D effect size was estimated and P-values were corrected for each feature to account for multiple comparisons using the Benjamini-Yekutieli procedure. Bootstrapped (1000 iterations) 95 % confidence intervals are presented. **HC:** Healthy Controls; **FTD:** Frontotemporal Dementia; **AD:** Alzheimer's Disease; **PD:** Parkinson's Disease; **LBD:** Lewy Body Dementia (comprising dementia in PD and Dementia with Lewy Bodies – DLB); **MCI-LBD:** Mild Cognitive Impairment in Lewy Body Dementia (comprising MCI in PD and MCI with reported Lewy Bodies pathology); **MCI-AD:** Mild Cognitive Impairment with reported AD pathology (or without Lewy Bodies).

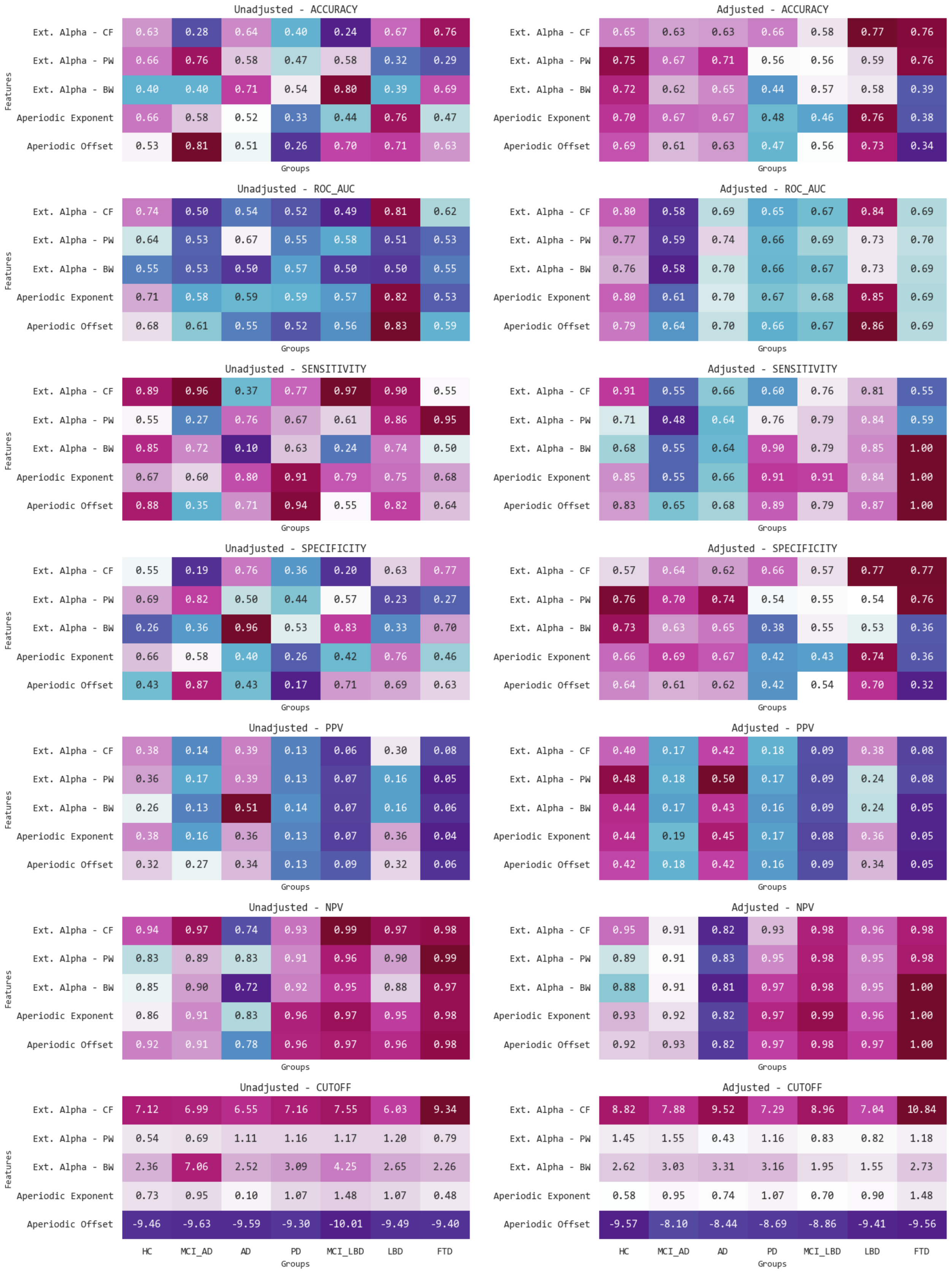


**Supplementary Figure 11. Discriminatory ability of Spectral Parameters as predictors of neurodegenerative diseases (NDDs).** Results from unadjusted (left column) and Age+Sex-adjusted (right column) multinomial logistic regression models with spectral parameters as predictors of NDDs. Heatmaps depict multiple performance metrics. **HC:** Healthy Controls; **FTD:** Frontotemporal Dementia; **AD:** Alzheimer's Disease; **PD:** Parkinson's Disease; **LBD:** Lewy Body Dementia (comprising dementia in PD and Dementia with Lewy Bodies – DLB); **MCI-LBD:** Mild Cognitive Impairment in Lewy Body Dementia (comprising MCI in PD and MCI with reported Lewy Bodies pathology); **ROC:** Receiver Operating Characteristics curve; **AUC:** Area Under the ROC curve; **PPV:** Positive Predictive Value; **NPV:** Negative Predictive Values; **Cutoff:** Cutoff obtained from Youden Index.
